# Breastfeeding limits the adverse impact of socioeconomic status on child health by modifying the infant gut microbiome

**DOI:** 10.1101/2025.06.23.25330007

**Authors:** Darlene L.Y. Dai, Melissa B. Manus, Courtney Hoskinson, Jie Jiang, Hind Sbihi, Kozeta Miliku, Susan C. Campisi, Daphne J. Korczak, Qingling Duan, Theo J. Moraes, Piushkumar J. Mandhane, B. Brett Finlay, Elinor Simons, Hannah Lishman, David M. Patrick, Padmaja Subbarao, Meghan B. Azad, Bo Chawes, Klaus Bønnelykke, Søren Johannes Sørensen, Jonathan Thorsen, Jakob Stokholm, Charisse Petersen, Stuart E. Turvey

## Abstract

Lower familial socioeconomic status (SES) is linked to increased childhood disease risk. Since SES has no inherent biological basis, identifying how it becomes physiologically embedded is essential for equitable intervention. Using data from the CHILD cohort, we analyzed modifiable pathways linking SES to child health and found that the infant gut microbiota plays a key mediating role. Breastfeeding was associated with a stabilized infant microbiota, buffering against environmental impacts and reducing health risks in lower SES contexts. The presence of *Bifidobacterium infantis*, enriched through breastfeeding, was linked to protection against adverse outcomes from SES. We observed similar associations in the independent COPSAC_2010_ cohort, including links among SES, breastfeeding, child health, the microbiota, and *B. infantis*. Together, these results suggest the improving breastfeeding rates and restoring breastfeeding-enriched microbes, like *B. infantis*, may help buffer early biological impacts of social inequality and support healthier trajectories for children growing up in industrialized settings.

**One-Sentence Summary:** Breastfeeding limits the adverse impact of socioeconomic status on child risk factors associated with non-communicable diseases potentially by modifying infant gut microbiota and enriching for *Bifidobacterium infantis*.

## Introduction

Socioeconomic disparity and health inequity are closely intertwined, with consequences that can span generations. Lower socioeconomic status (SES), a social construct with no inherent biological basis, is associated with heightened risk of chronic non-communicable diseases (NCDs), including cardiometabolic disease, chronic respiratory disease, and mental illness^1–4^. These health disparities can emerge early in life, as limited socioeconomic resources increase children’s likelihood of developing NCD-associated conditions^3,5,6^. Moreover, industrialization has coincided with an increased prevalence of NCDs, which globally now account for approximately 70% of all deaths, exacerbating both the issue and its disproportionate impact on vulnerable populations^7,8^. Understanding how socioeconomic inequity becomes biologically embedded is therefore essential, yet the mechanisms driving this process remain unclear. Identifying modifiable pathways through which SES affects health could inform innovative strategies that equitably support childhood health and ultimately reduce the burden of NCDs^9,10^.

The rise of NCDs closely follows factors linked to industrialization, such as antibiotic use, urbanization, processed food diets, and a highly sanitized environment, all of which have reshaped human interactions with the microbial world^11–13^. As a result, industrialization has driven intergenerational changes in both our individual microbiome, the communities of microbes living in and on our bodies, as well as the broader metacommunity of microbes capable of colonizing all of us^14,15^. Researchers have justifiably identified the microbiota as a potential pathway by which essential microbial exposures that support healthy development are diminished or altered^8,16,17^. Many SES-related factors also affect the microbiota, especially during early life, including breastfeeding initiation and duration, antibiotic exposure, and access to green spaces and fresh foods^10,18–20^. Since the infant microbiota is both highly sensitive to disruptions and critical for healthy development, it may serve as a nexus linking SES-related exposures to childhood health outcomes. Thus, safeguarding or restoring the early-life microbiota represents an innovative opportunity to limit the rise of NCDs^18^. However, measures to ensure early exposure to essential microbes, while limiting factors that disrupt health-promoting microbial development, must be implemented equitably to help break cycles of health disparities^9^.

The infant microbiota matures alongside multiple rapidly developing physiological systems, such as the immune, metabolic, and neurobehavioral systems^21–23^. Early-life microbiota disruptions have been associated with multiple diseases linked to metabolic dysfunction, allergies, and chronic inflammation^24–29^. While a growing body of evidence suggests that familial SES influences the infant and childhood gut microbiota^30–34^, comprehensive analyses of the environmental factors mediating this relationship are limited. Moreover, SES is a multidimensional construct that typically encompasses not only family income but also educational attainment and subjective perceptions of social status, yet studies typically investigate these different social factors in isolation^30–32^. Integrating SES parameters with early-life exposures to define their impact on the infant microbiota and health could identify critical SES-related drivers of microbiota development, highlighting opportunities to optimize the early-life microbiota and improve population health.

In this study, we leveraged the CHILD Cohort Study, the largest prospective birth cohort in Canada, which includes diverse families across socioeconomic, geographic, and ethnic backgrounds. The study captured perinatal measures of SES, parental demographics and history, environmental exposures, infant gut microbiota profiles, and physician-defined health outcomes at age five^35^. We investigated how familial SES shaped early-life exposures and parental well-being and assessed its associations with infant microbiota composition and later child health. Regression models revealed considerable overlap between the associations of familial SES and breastfeeding rates on both infant microbiota composition and multiple health outcomes, with consistent directional patterns. Furthermore, microbes enriched by breastfeeding - most notably *Bifidobacterium infantis* - were linked to protection against several childhood health risk factors for NCDs. We replicated key associations in the independent COPSAC_2010_ cohort, reinforcing links among SES, breastfeeding, the infant microbiome, and child health. Together, these findings suggest that supplementation with key beneficial bacteria, alongside efforts to reduce breastfeeding barriers, may offer scalable strategies to improve child health and reduce socioeconomic health disparities.

## Results

### Familial SES is associated with multiple domains of perinatal experience and links to future adverse child health outcomes

The CHILD Study collected five measures of SES when enrolling pregnant women and their partners in the study at approximately 18 weeks of gestation. These included household income, highest education of mother and father, and the MacArthur scale of subjective social status in both Canada and their local community^35,36^. Of the 3,263 eligible families enrolled in the general cohort of the CHILD study (Extended Data Fig. 1a, Extended Data Table 1), 2,752 children had complete data of the five individual SES factors and were used to construct a single ’SES index’ integrating all five measures based on confirmatory factor analysis (CFA)(Fig. 1a). Independent cluster analysis confirmed the index’s internal consistency (⍺ = 0.76) (Extended Data Fig. 1b). A single SES index was derived from this estimated latent factor and was used in all downstream analyses.

**Fig. 1.**
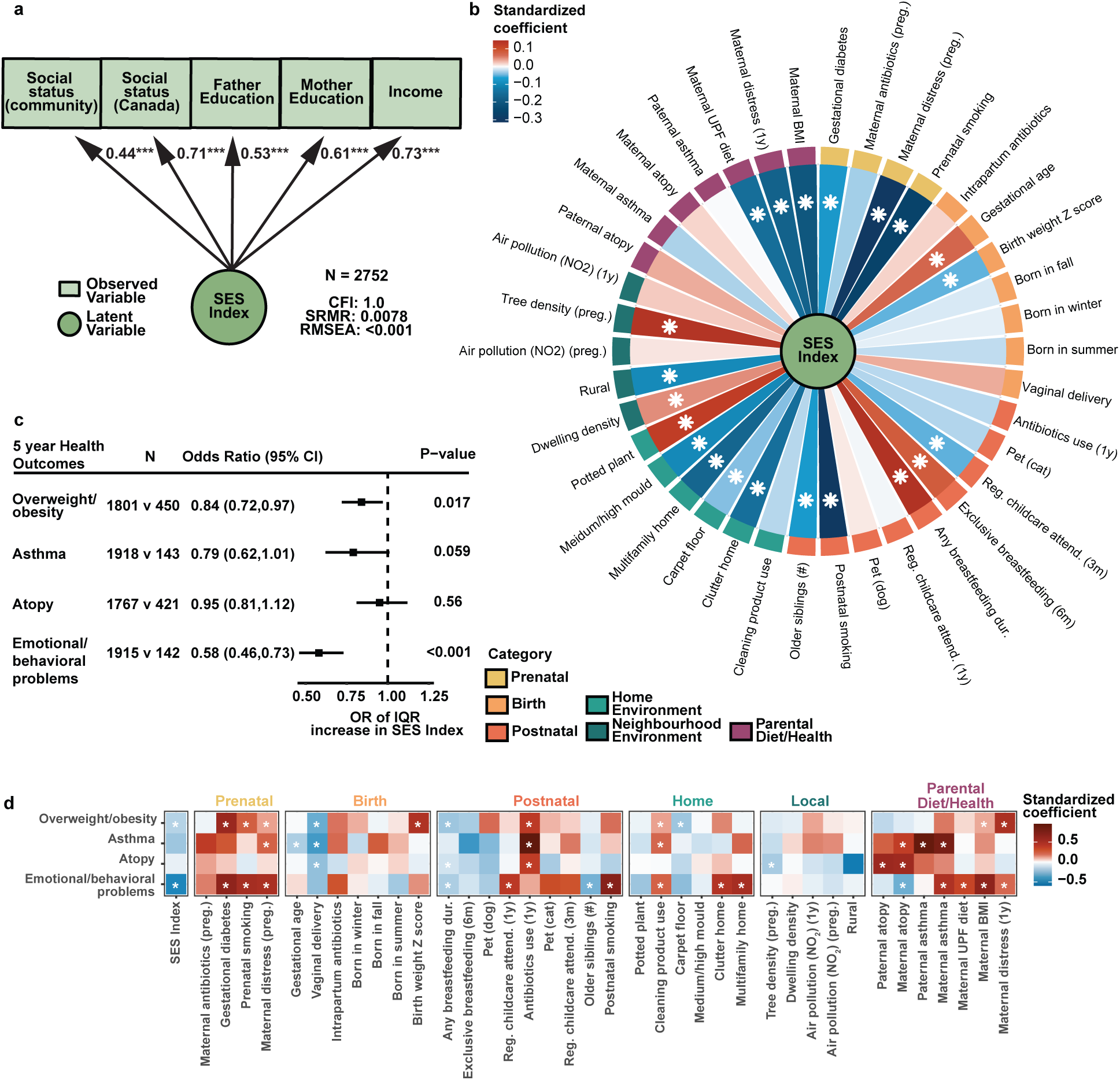
Summarized SES index is associated with childhood NCDs and perinatal factors and exposomes. (a) The summarized SES index is generated using observed SES factors based on confirmatory factor analysis. In the figure, circles represent unobserved latent variables and rectangles represent observed variables. One-headed arrows represent the correlations between two variables and the standardized coefficient is shown. * p<0.05, ** p<0.01, and *** p<0.001. CFI, comparative fit index; RMSEA, root mean square error of approximation; SRMR, standardized root mean residuals. (b) Associations between SES and perinatal factors and the broad exposome. Data were presented using standardized coefficient of perinatal factors on summarized SES index during pregnancy. Red indicates positive association, whereas blue indicates negative association. Stars represent significant associations (p < 0.05) using least square regression models. (c) Forest plot of odds ratio (95% CI) of SES index on childhood NCDs at 5 years of age. Data are presented as odds ratio of interquartile change of SES index ± standard error of the mean. (d) Heatmap of the associations between childhood health outcomes at 5 years of age and perinatal factors and exposomes. Associations were measured using standardized coefficients of logistic regressions with adjustment of study site and sex. Red represents positive association and blue represents negative association. Stars represent significant associations (p < 0.05) using least square regression models.

We next sought to understand how SES related to nearly 40 perinatal factors collected between 18 weeks of gestation and infants’ 1-year visit. These factors spanned parental diet and health, prenatal exposures, specific measures related to birth, postnatal exposures within the first year, and the home or local community environment. The summarized SES index was compared to each factor using regression models with adjustment of study center and sex assigned at birth (Fig. 1b). Of the 36 perinatal variables analyzed, 21 (58.3%) were associated with SES (p-value<0.05). Moreover, SES was significantly linked to factors across multiple exposure types, highlighting the broad and integral role of SES in shaping parental health and exposures during the perinatal period. To confirm the robustness of our findings, we applied multiple sensitivity analyses. These included a comparison of our prenatal SES index to one created using data collected at 1 year; a sensitivity analysis adjusting for genetic ancestry of participants; a sub-cohort analysis within each study center; and sensitivity analyses using individual observed SES factors, all of which indicated that our SES index is a good representor of the 5 observed SES factors and robustly associates with perinatal factors (Extended Data Fig. 1c).

Socioeconomic status has furthermore been associated with the likelihood of developing NCDs^3,5,6^. We therefore compared adverse health outcomes collected at the 5-year visit in CHILD considered to be early risk factors for multiple NCDs, including atopic (aka. allergic) sensitization (defined by a positive allergen skin prick test), physician diagnosis of asthma, overweight or obesity classifications (determined by BMI z-scores), and emotional/behavioral problems (defined using the childhood behavioral checklist). Infants from families with higher SES had significantly lower odds of overweight/obesity (Odds ratio (OR) for an interquartile (IQR) increase in SES index=0.84, p=0.017) and emotional or behavioral problems (OR=0.58, p<0.001), with a trend towards protection from asthma (OR=0.79, p=0.059). In contrast, SES was not associated with 5-year atopic sensitization (Fig. 1c). Reinforcing the importance of the early life environment in the developmental origins of health and disease, 23 of the 36 perinatal factors were significantly (p-value<0.05) associated with at least one 5-year health outcome (Fig. 1d). Moreover, some were consistently associated with at least three distinct health outcomes when accounting for both significance and directionality. Notable protective factors included longer breastfeeding duration and vaginal delivery, which were negatively associated with childhood risk factors. In contrast, frequent use of cleaning products in the home, early antibiotic exposure (within the first year), and maternal distress during pregnancy were positively associated with three or more adverse health outcomes, indicating their relevance for early childhood development. Together these data demonstrate that SES is linked to risk factors for multiple distinct NCDs within the CHILD cohort, supporting further investigating the biological underpinnings of SES.

### Familial SES significantly influences the infant gut microbiota

The infant gut microbiota is highly sensitive to disruption, including by factors associated with SES, and changes in the microbiota during early life can have long-lasting effects on physiological and behavioral development^24,28,29,37,38^. After defining the associations between SES and perinatal exposures (Fig. 1), we next examined how SES and these perinatal factors were associated with the infant microbiota. We analyzed shotgun metagenomic profiles of the gut microbiota from 1,479 participants with stool samples collected at clinical assessment scheduled for 3 months and/or 1 year of age (Extended Data Fig. 1). Using both Wilcoxon tests (comparing SES quartiles) and linear mixed-effect models (assessing SES as a continuous variable), we examined associations between SES and species alpha diversity, measured by the Shannon index, at 3 months, 1 year and its change by 1 year, adjusting for technical covariates and study site (Fig. 2a, Extended Data Fig. 2a). Infants from higher-SES families had significantly lower alpha diversity at 3 months (β_SES_=– 0.045, p=0.0053; Wilcoxon p<0.001) but showed a greater increase in diversity over time (β_SES_=0.065, p=0.0015; Wilcoxon p=0.0056). Although higher microbial biodiversity is typically associated with positive health outcomes, there exists a well-known paradox that breastfeeding, considered beneficial early in life, constrains or delays diversification in early infancy^24,39^. Consistent with this, breastfeeding rates at 3 months and 1 year were higher in the highest SES quartile (Q4) (89% and 45%, respectively) than in the lowest SES quartile (Q1) (71% and 31%). Notably, when adjusting for current breastfeeding, the differences in alpha diversity between lower and higher SES groups became non-significant (Extended Data Fig. 2a), suggesting that the observed associations were driven by higher breastfeeding rates in families with higher SES. We then evaluated the association between SES and the taxonomic microbiota composition (beta diversity) at 3 months and 1 year using Euclidean distances (Fig. 2b). Principal coordinate analysis (PCoA) revealed significant differences between the lowest and highest SES quartiles at 3 months along the first two PCoA axes (p<0.001), with a clear dose-response associations (PCoA1 β_SES_=1.62, PCoA2 β_SES_=-1.57, p<0.001). Within one-year samples, SES associations were more modest but still evident in the third PCoA axis, which was significantly associated with the SES index (PCoA3 β_SES_=-0.70, p=0.013) (Fig. 2b, Fig S2b). These results indicate that the association between SES and the infant microbiota is strongest at 3 months but still detectable at 1 year.

**Fig. 2.**
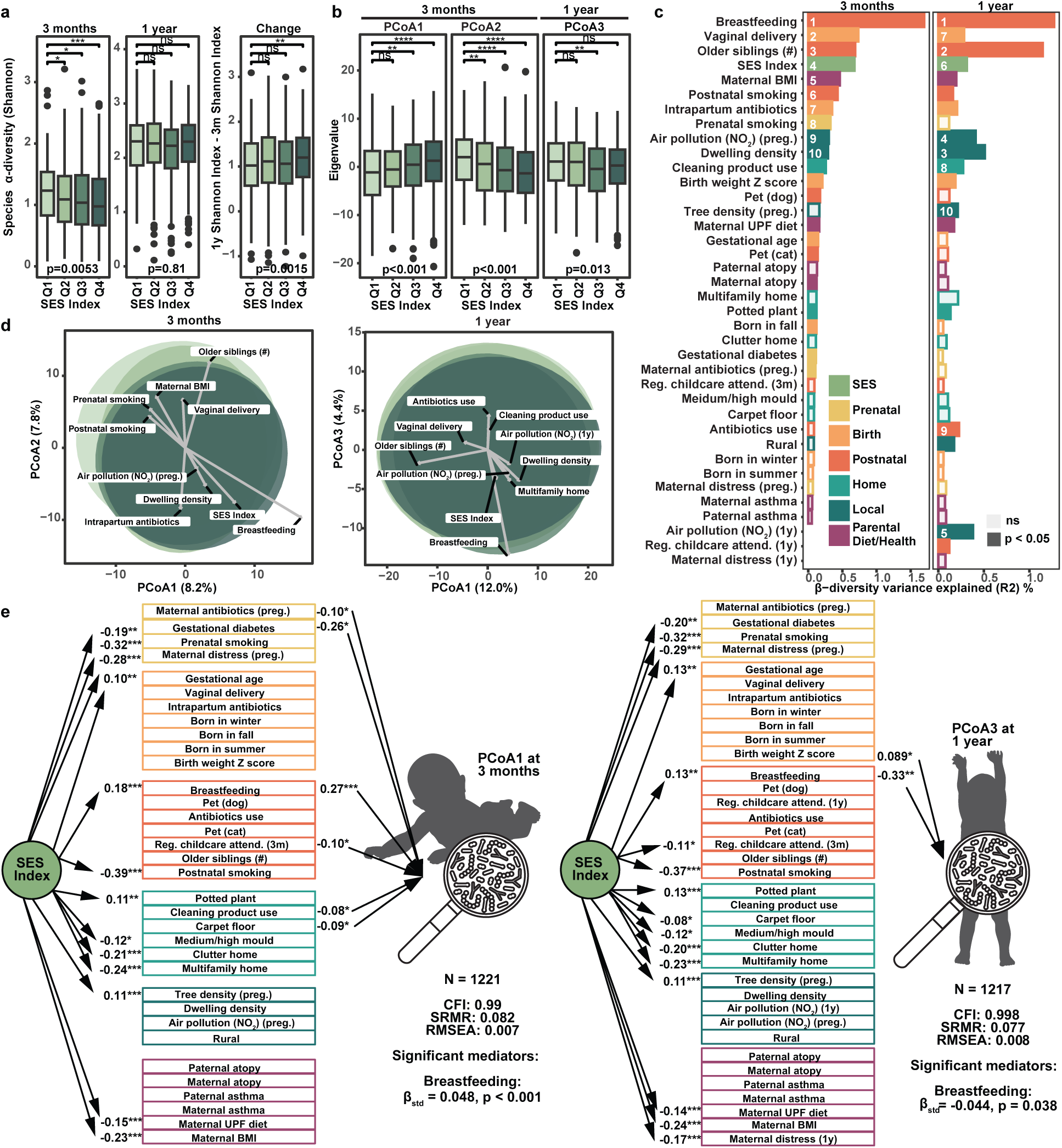
SES a top influencer on infant gut microbiota, primarily through breastfeeding. (a) Differences in gut microbiota species α-diversity (Shannon index) at 3 months, 1 year, and change from 3 months to 1 year, across children with different quartiles of SES index. Specifically, SES Q_1_ includes samples with the lowest 25% SES index and SES Q_4_ includes samples with the top 25% SES index. Differences across SES quartiles compared to the first quartile of SES were estimated based on Wilcoxon test. P-value at the bottom of each figure was based on a linear least squares regression model on the continuous SES index. (b) Boxplot of the significant top 3 PCoAs across SES quartiles. Differences across SES quartiles compared to the first quartile of SES were estimated based on Wilcoxon test. P-values at the bottom of each figure were based on least-squares linear regression models using the continuous SES index. ns p>0.05, * p<0.05, ** p<0.01, and *** p<0.001. (c) Barplots of beta-diversity variance explained (R^2^) by each individual factor for infant gut microbiota at 3 months and 1 year, and the top 10 significant factors ranked by R^2^ are marked. Only factors with a p-value < 0.05 are filled with color, which indicates the categories of these factors. (d) Principal coordinate analysis (PCoA) visualizing the beta-diversity of the 3-month and 1-year infant gut microbiota. Colour indicates the quartiles of SES index. The arrows represent the strength and direction of certain metadata variables that explain microbiome variation using Pearson correlation. (e) Visualization of SEM model testing the causal relationship between SES index, perinatal factors and exposomes, and PCoA1 of infant gut microbiota at 3 months (left) and PCoA3 of infant gut microbiota at 1 year (right). The mediating effects of perinatal factors and exposomes were tested using SEM model and only significant indirect effects and associations (one-head arrows) (p < 0.05) are shown in the figures.

As SES itself is a social construct that is not directly biologically linked to the microbiota, we explored which perinatal exposures could be mediating this effect. For each timepoint, we applied permutation-based multivariate analysis of variance (PERMANOVA) with the study center as strata and adjusting for technical covariates, to compare the influence of SES and each perinatal factor (Fig. 2c). SES ranked as the fourth most influential factor at 3 months (R²=0.68%, p<0.001) and sixth at 1 year (R²=0.31%, p<0.001). Only extensively studied influencers of the infant microbiota, such as breastfeeding status, delivery mode and the number of older siblings, had a stronger association than SES at 3 months (Fig. 2c). A biplot visualizing the top 10 influencing factors at both timepoints indicated that SES and breastfeeding consistently moved in similar directions at both timepoints (Fig. 2d). To further investigate the mediating role of breastfeeding, we employed structural equation modeling (SEM), a statistical approach that tests how well observed data fit a hypothesized causal framework. Using PCoA1 at 3 months and PCoA3 at 1 year to represent the overall SES-associated gut microbiota, we found that among the 36 perinatal factors, current breastfeeding at the time of stool sample collection was the only significant mediator at both 3 months (indirect effect β_std_=0.048, p<0.001) and 1 year (β_std_=-0.044, p=0.038) (Fig. 2e). These findings indicate that differences in breastfeeding mediated the relationship between SES and the microbiota at both timepoints, suggesting that breastfeeding may be a key pathway through which SES-related health inequities arise within the infant microbiota to impact long-term health trajectories.

### Breastfeeding shields the infant microbiota from disruption and can mitigate health risks in infants born to lower SES families

Human milk plays a key role in shaping the infant gut microbiota and has been demonstrated to pace microbial development and select for keystone species critical for infant health^39–41^. We previously found that breastfeeding attenuated the impact of early antibiotic exposure by limiting microbiota disruption and reducing subsequent asthma risk ^24^. To assess the broader extent of this buffering effect, we first compared the overall beta-dispersion of 3-month samples and 1-year samples based on participant breastfeeding status (Fig. 3a). Participants who either never breastfed or were no longer breastfeeding (No BF) at 3 months and the time of the stool sample collection visit were compared to infants who were still breastfeeding (BF) at the time of sample collection. At 3 months, the BF infants had significantly lower beta-dispersion compared to the No BF infants (p-value<0.001), in agreement with the constraining properties of human milk on the infant microbiota (Fig. 3a). These differences in beta-dispersion were no longer detectable in the 1-year stool sample (p-value=0.6), by which point all infants were consuming a variety of complementary foods, thus diminishing the ‘constraining’ influence of human milk. We next reran our PERMANOVA analysis of perinatal factors within each subgroup, using a bootstrapping method to account for differences in sample size. The mean explained variance across 100 runs was compared between BF and No BF infants, and variables with p<0.05 in 80% or more of runs were considered significant. At 3 months, 20 perinatal factors were significantly associated with microbiota composition in No BF infants, whereas only three factors remained significant in BF infants (Fig. 3b). Similarly, at 1 year, although breastfeeding no longer significantly constrained microbiota beta-dispersion, only 1 of the 18 perinatal factors that explained significant variance in No BF infants remained significant in BF infants. These data suggest that the presence of human milk is associated with lower microbiota variability in young infants, as measured by beta-dispersion, and with reduced influence of external factors during the first year of life, consistent with a potential shielding effect of breastfeeding against microbiota disruption.

**Fig. 3.**
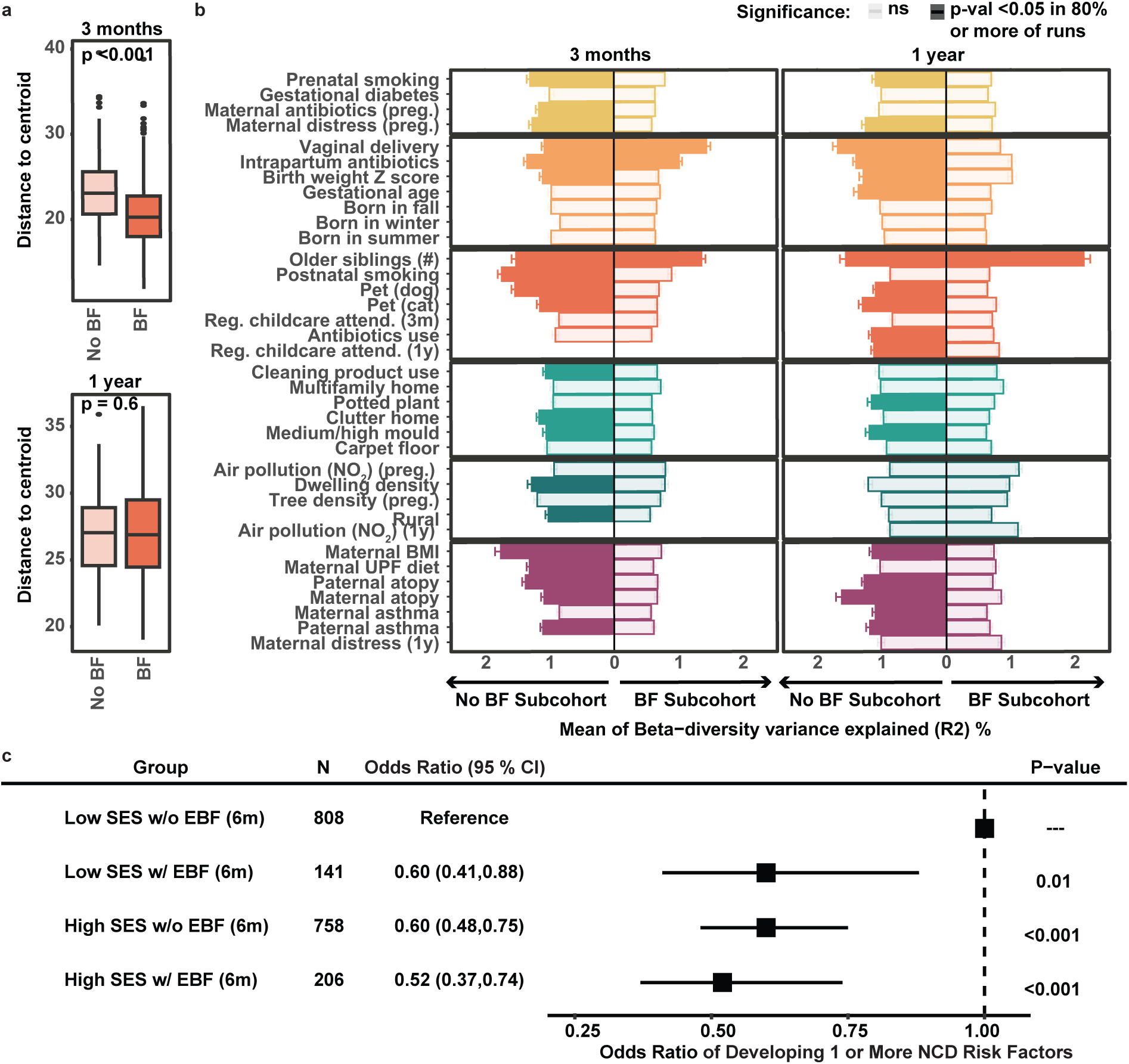
Breastfeeding shields the infant microbiota from disruption and can mitigate health risks in infants with limited SES. (a) Boxplot of beta-dispersion at 3 months and 1 year for children who were still breastfed (BF) at the stool sample collection age (3 months N=159, 1 year N=149) and children who either never breastfed or were no longer breastfeeding (No BF) by 3 months and the stool sample collection age (3 months N=1001, 1 year N=479). Beta-dispersion is calculated based on Euclidean distance using modified centered log-ratio (mCLR) transformed relative abundance of species. P-values of the difference were estimated based on Wilcoxon test. (b) Barplots of beta-diversity variance explained (R^2^) by each factor for infant gut microbiota at 3 months and 1 year in subgroup BF and No BF infants using bootstrapping method (100 runs with random sampling with replacement using the same sample size). Error bars for significant factors present ± standard error of the mean around the mean of R2 across 100 bootstrapping runs. Variables with p<0.05 for at least 80% of runs were considered significant and filled with colors, which indicate the categories of these factors. (c) Forest plot of odds ratio (95% CI) of compound SES and exclusive breastfeeding status (EBF) at 6 months on one or more SES-associated NCDs (childhood overweight/obesity, asthma and emotional/behavioral problems at 5 years of age).

To assess the association between breastfeeding and child health outcomes, we divided participants into above and below median of the SES index and quantified their likelihood of developing one or more of the SES-associated NCD risk factors (childhood overweight/obesity, asthma or emotional/behavioral problems at 5 years) based on whether they were exclusively breastfed for six months, which is the duration currently recommended by the WHO. Among participants from low-SES families, exclusive breastfeeding for six months was associated with an approximately 40% reduced odds of developing one or more NCD risk factors (OR=0.60, 95%CI 0.41-0.88, p = 0.01) (Fig. 3c). Notably, this protective association was comparable to that observed in participants from high-SES families, who were protected compared to low-SES families regardless of breastfeeding status (OR=0.60, 95%CI 0.48-0.75 and OR=0.52, 95%CI 0.37-0.74 for high-SES without and with BF, respectively, both p<0.001). These findings suggest that breastfeeding not only mediates the effect of SES on the infant microbiota but also mitigates microbial disruptions and reduces adverse health outcomes among children from higher-risk, lower-SES families.

### Replication in the independent COPSAC_2010_ Cohort

To validate our findings in an independent cohort, we turned to the Copenhagen Prospective Studies on Asthma in Childhood 2010 (COPSAC_2010_), a population-based Danish birth cohort comprising 700 mother-child pairs. SES was assessed at enrollment of the child 1 week after birth, using three indicators: household income and maternal and paternal education in the pregnancy period. Among 681 children with complete SES data, a latent SES index was constructed (Fig. 4a). Consistent with results from the CHILD cohort, higher SES in COPSAC_2010_ was associated with reduced risk of physician-diagnosed asthma at age five (OR per IQR increase in SES index = 0.73, p = 0.03), emotional or behavioral problems (OR=0.45, p<0.001) at age six and a trend towards protection from overweight/obesity at age five (OR=0.71, p=0.078). No association was observed between SES and atopic sensitization at age six (Fig. 4b).

**Fig. 4.**
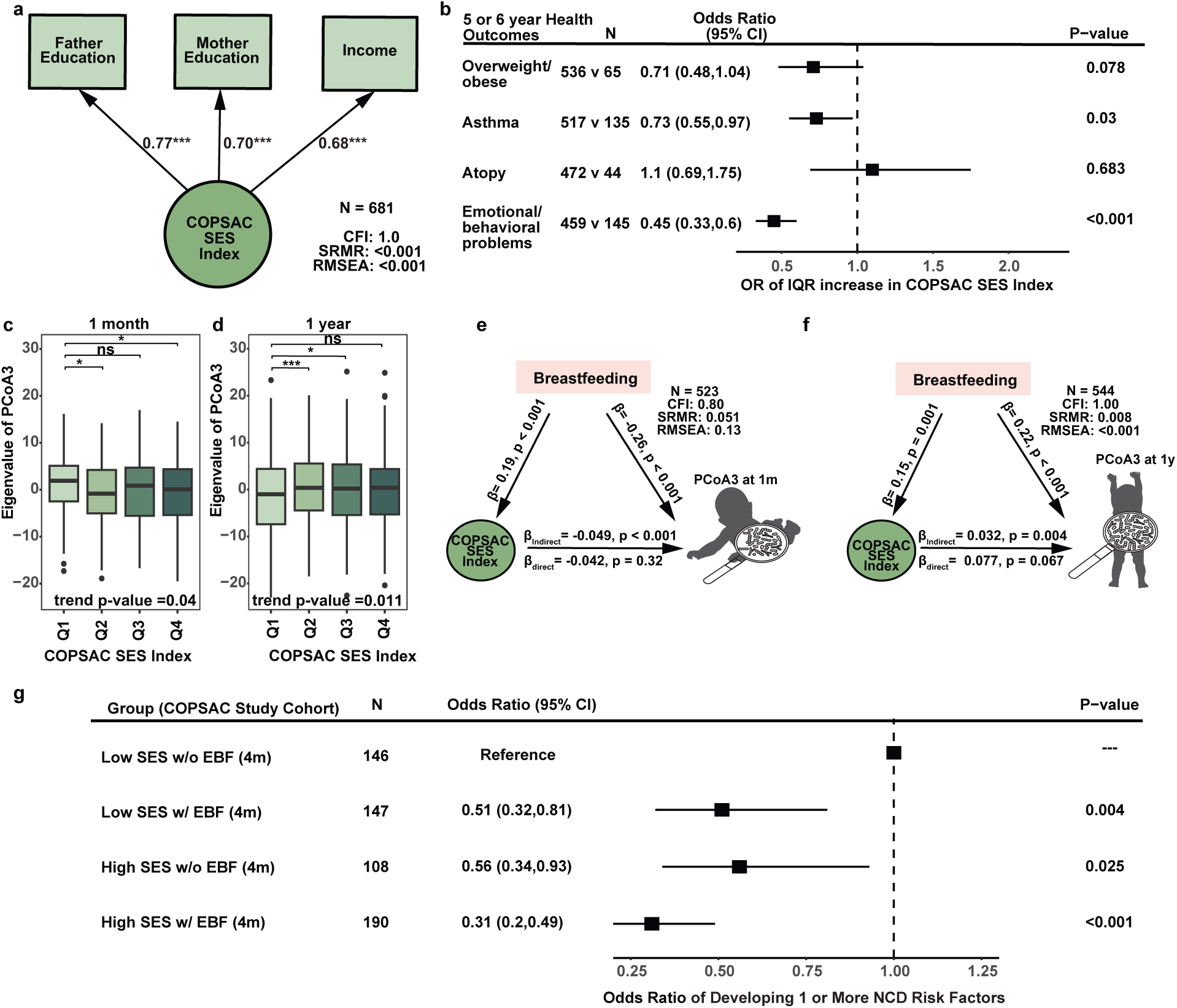
Replication on the independent COPSAC_2010_ cohort. (a) The summarized COPSAC_2010_ SES index is generated using observed SES factors in COPSAC_2010_ cohort based on confirmatory factor analysis. In the figure, circles represent unobserved latent variables and rectangles represent observed variables. One-headed arrows represent the correlations between two variables, and the standardized coefficient is shown. * p<0.05, ** p<0.01, and *** p<0.001. CFI, comparative fit index; RMSEA, root mean square error of approximation; SRMR, standardized root mean residuals. (b) Forest plot of odds ratio (95% CI) of COPSAC_2010_ SES index on childhood NCDs at 5 years of age in COPSAC_2010_ cohort. Data are presented as an odds ratio of the interquartile change of SES index ± standard error of the mean. (c-d) Boxplot of the PCoA3 across SES quartiles at 1 month (c) and 1 year (d). Differences across SES quartiles compared to the first quartile of SES were estimated based on Wilcoxon test. P-values of trend at the bottom of each figure were based on least-squares linear regression models using a continuous SES index. ns p>0.05, * p<0.05, ** p<0.01, and *** p<0.001. (e-f) Visualization of SEM model testing the causal relationship between SES index, breastfeeding, and PCoA3 of infant gut microbiota at 1 month (e) and 1 year (f). The mediating effects of breastfeeding were tested using an SEM model. (g) Forest plot of odds ratio (95% CI) of compound COPSAC_2010_ SES and exclusive breastfeeding status at 4 months on one or more SES-associated NCDs (childhood overweight/obesity and asthma at 5 years of age).

We next tested whether breastfeeding mediated the relationship between SES and the infant gut microbiome in the COPSAC_2010_ study using SEM. Shotgun metagenomic sequencing data were available for 523 subjects at the 1-month visit and 544 subjects at the 1-year visit. Principal coordinate analysis was applied to gut microbiota profiles at 1 month and 1 year. In contrast to the CHILD cohort, SES-related variation in COPSAC_2010_ microbiota profiles was limited to PCoA3 at both time points (1-month p=0.04; 1-year p=0.011) (Fig. 4c–d), possibly reflecting reduced SES variability within the cohort. These axes were used in SEM to capture SES-associated microbiota variation. At 1 year, breastfeeding showed a significant indirect effect (β=0.032, p=0.004). Although the SEM model at 1 month exhibited limited fit, possibly due to reduced variation or sample size constraints, the indirect effect of breastfeeding remained statistically significant (β=- 0.049, p<0.001). Together with the CHILD results, these findings support a consistent role for breastfeeding in shaping SES-related microbiota trajectories.

Finally, we assessed whether breastfeeding modified SES-related health outcomes in the COPSAC_2010_ cohort. Children were stratified by SES index (lower vs. upper half), and exclusive breastfeeding was evaluated in relation to risk of asthma, overweight/obesity and emotional/behavioral problems. Although the WHO recommends six months of exclusive breastfeeding, we focused on four months, as this aligned with the introduction of solid foods for many infants in the cohort, resulting in few mothers exclusively breastfeeding to six months. Among children from lower-SES families, exclusive breastfeeding for four months was associated with a 39% reduction in the odds of adverse outcomes (OR=0.51, 95% CI: 0.32–0.81, p=0.004) (Fig. 4g). These findings mirror those from the CHILD cohort and highlight breastfeeding as a modifiable factor that may help mitigate the effects of socioeconomic disadvantage on early microbiota development and long-term health risk.

### Most SES-associated species are associated with breastfeeding in consistent directions

Within our own CHILD Study Cohort, we next identified specific microbial species that may be impacted by SES. We identified these species using longitudinal analyses to capture both temporal changes and overall differences within the microbiota in the first year of life. Using linear mixed-effect models (MaAsLin2), we identified 30 species significantly associated with SES, either through differences in overall abundance or changes over time (FDR<0.1). Of these, 5 species showed overall higher colonization with increasing SES, including *Bifidobacterium infantis*, *Phocaeicola dorei*, *Bacteroides fragilis*, *Bacteroides thetaiotaomicron,* and *Bacteroides caccae*. Conversely, 3 species showed consistently lower colonization with higher SES, including *Ruthenibacterium lactatiformans*, *Clostridium innocuum*, and *Phocaeicola vulgatus* (Fig. 5a, Extended Data Fig. 3a). These 8 species exhibited relatively stable SES-associated differences across infancy. In contrast, 22 species showed significant differences in temporal changes in colonization based on familial SES (Fig. 5a, Extended Data Fig. 3a), highlighting SES’s extended association with microbial composition within the first year of life. Notably, all 30 species were associated with at least one of the 36 perinatal variables using similar models, with breastfeeding status linked to 23 (77%) of the species in a consistent direction (Fig. 5a, Extended Data Fig. 3b).

**Fig. 5.**
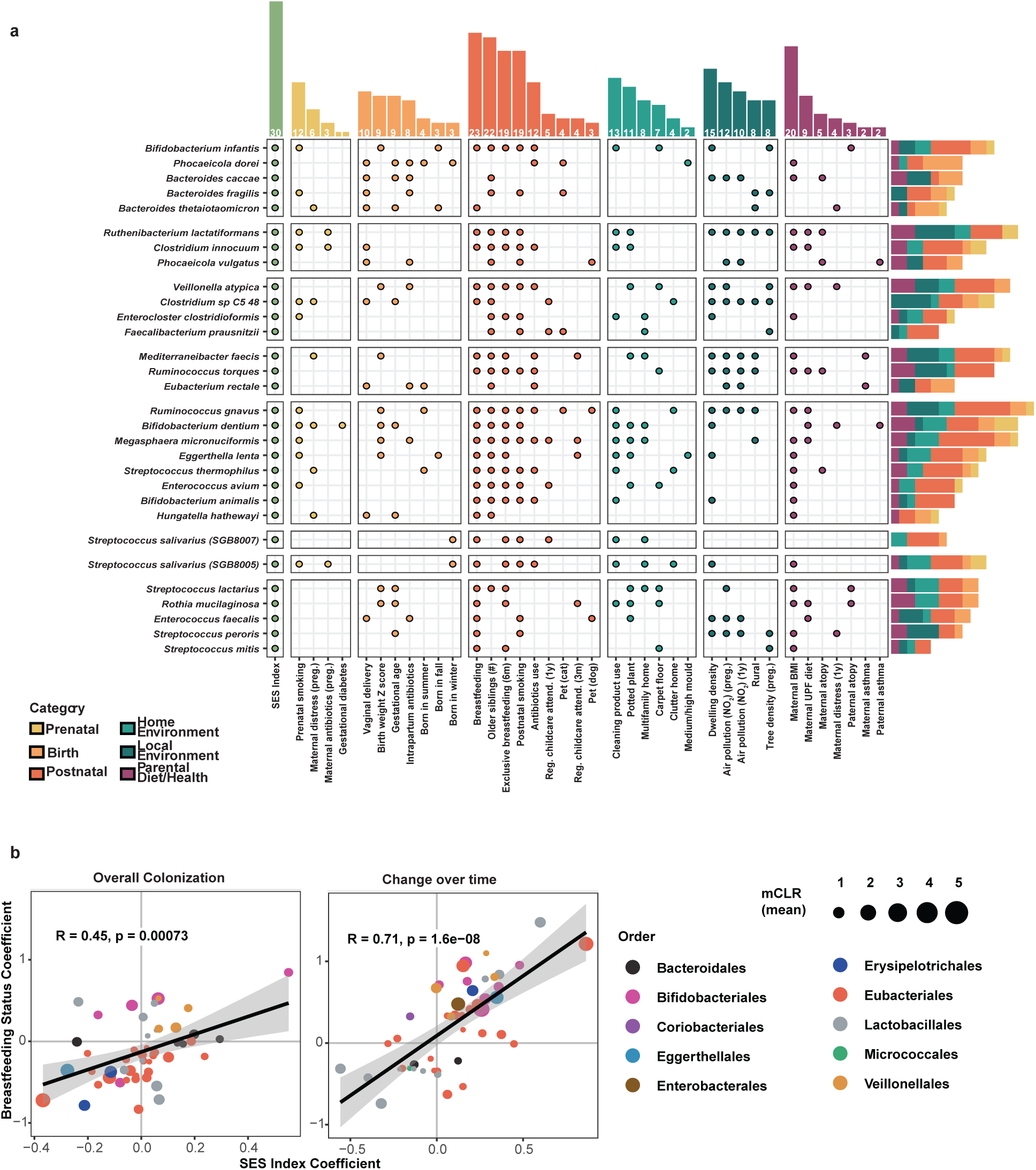
Most of the SES-associated species are associated with breastfeeding and other perinatal factors and exposomes. (a) Associations between perinatal factors and SES based on MaAslin2 models for SES-associated species. Dots represent the significance (FDR < 0.1) in either slope or overall effect (methods) and their colors represent the categories of factors. The top bar represents the number of significant species for each factor among the 30 SES-associated species. The right-side bar represents the number of significant factors for each species. Species are grouped based on the pattern of change across time and socioeconomic status shown in Extended Data Fig. 3a. Child antibiotic use was defined as antibiotic exposure before the stool sample collection age, and breastfeeding was defined as children who were still breastfed at the stool sample collection age. (b) Correlation plot of overall effect (MaAslin2 models without interaction measuring overall colonization) and slope effect (MaAslin2 models with interaction measuring change over time) between SES and breastfeeding for species significantly associated with either SES or breastfeeding (FDR of slope or overall effect < 0.1). Dot color represents the species family and size represents the mean of the relative abundance of species across 3 months and 1 year.

This was further demonstrated by comparing the coefficients for overall species colonization and changes in species abundance over time when testing either the SES index or breastfeeding status in our longitudinal analyses. Notably, for microbes significantly associated with SES or breastfeeding, both the overall effect (overall colonization levels, 53 species) and the slope effect (colonization changes over time, 48 species) showed a strong positive correlation between SES and breastfeeding coefficients (overall effect: Pearson R=0.45, p=0.0007; slope effect: R=0.71, p=1.6e-08) (Fig. 5b). Moreover, functional characterization of the shotgun sequencing profiles using MetaCyc pathway classifications revealed a similar relationship between SES and breastfeeding. Of a total of 374 pathways presented in at least 10% of samples, 150 (40%) were linked to familial SES with similar correlations detected in both overall colonization and changes over time between SES and breastfeeding status (Pearson R=0.45 and 0.75, respectively) (Extended Data Fig. 3c). These data reinforce the interconnected influence of SES and breastfeeding on shaping both the taxonomic composition and functional potential of the infant microbiota.

To assess the reproducibility of these findings, we applied the same MaAsLin2 modeling approach to the COPSAC_2010_ cohort. Among the 151 microbial species detected in at least 10% of COPSAC_2010_ participants, 15 were significantly associated with SES (FDR < 0.1) (Extended Data Fig. 4a). Of these 15 species, 5 were also associated with breastfeeding, including *B. infantis*, *Escherichia coli*, *Veillonella atypica*, *Streptococcus salivarius* (SGB8005), and *Streptococcus parasanguinis*. Notably, four of these species overlapped with the CHILD cohort findings: *B. infantis*, *V. atypica, B. dentium*, and *S. salivarius* (SGB8005). Given the geographic and national differences between the COPSAC_2010_ and CHILD cohorts, variation in microbiota composition was expected; however, the overlap of four key species and 4 out of the 15 SES-associated species were also associated with breastfeeding in consistent direction reinforces the consistency of these associations across cohorts (Extended Data Fig. 4b).

### Health benefits afforded by familial SES are mediated through changes in the infant microbiota

Our findings so far indicate that familial SES is associated with both the infant gut microbiota and multiple distinct health outcomes at 5 years in two independent cohorts. To explore whether modifying the microbiota could improve childhood health, we examined whether the microbiota mediated associations between SES and health outcomes. We again used SEM, in which the microbiota was represented by a latent factor comprising the 30 SES-associated species abundances at 3 months and 1 year using confirmatory factor analysis. Consistent with the lack of association between SES and 5-year atopy, the indirect effect of the SES-associated microbiota on atopy was not significant (β_std_=0.003, p=0.19). In contrast, the SES-associated microbiota emerged as a significant mediator for childhood overweight/obesity (β_std_=-0.015, p=0.001), asthma (β_std_=-0.017, p=0.002), and emotional/behavioral problems (β_std_=-0.10, p<0.001) at 5 years of age (Fig. 6a). These results support the hypothesis that the infant microbiota plays a crucial role in linking familial SES to NCD-associated childhood outcomes and underscore the importance of identifying and supporting specific beneficial microbes to promote healthy child development. Of the 30 microbial species linked to familial SES, eight were associated with elevated weight gain, three with asthma, and two with emotional or behavioral problems at age five (Fig. 6b). Notably, *B. infantis* was the only species linked to two of the three SES-related outcomes, overweight and asthma, in a consistently protective direction (Fig. 6b, Extended Data Fig. 5). It also showed a significant protective association with atopy.

**Fig. 6.**
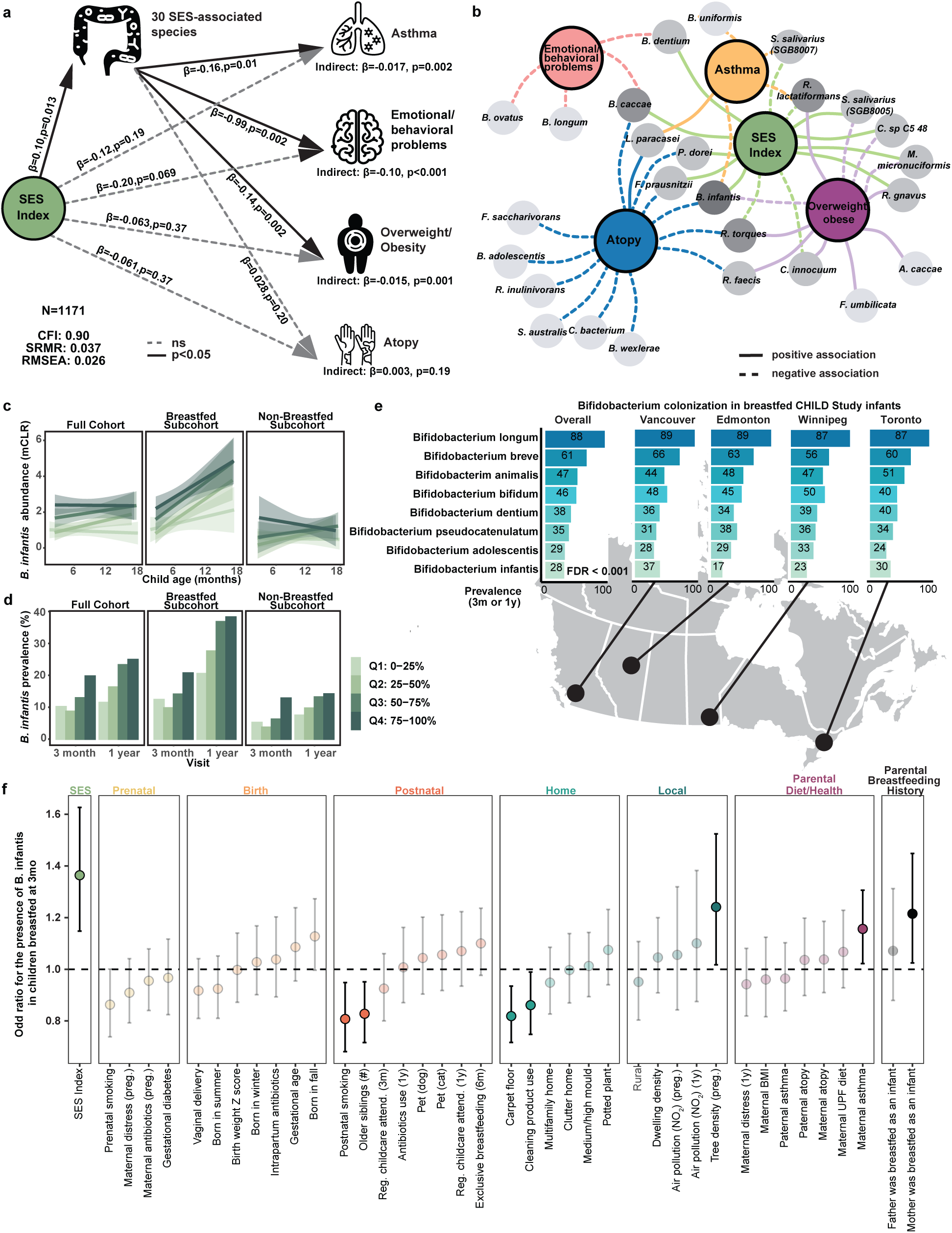
Higher family SES is associated with lower childhood overweight/obesity, asthma risk and emotional/behavioral problems by shaping the infant gut microbiota. (a) Visualization of SEM testing the mediation effect of SES-associated microbiome on childhood overweight/obesity, asthma, atopy and emotional/behavioral problems at 5 years of age. Infant gut microbiome was conceptualized to include the 30 SES-associated species at 3 months and 1 year. Significant associations (p < 0.05) are solid black lines and non-significant associations are gray dashed lines. All regressions in SEM models were adjusted for the sex, study site, stool sample collection age and processing time. (b) Species associated with any childhood NCDs (FDR < 0.1) based on MaAslin2 models with or without interaction. The association is based on the slope effect and the overall effect if the slope effect is not significant. The solid line indicates a significant positive association, and the dashed line indicates a significant negative association. The darkness of gray for each species is based on the number of associated phenotypes. (c) Relative abundance and (d) prevalence of *B. infantis* among all children, children with and without breastfeeding at the stool sample collection age. Colour indicates the quartiles of SES index. (e) Prevalence of *Bifidobacterium* species across CHILD study sites in Canada among children breastfed at 3 months. Only the prevalence of *Bifidobacterium infantis* was significantly different across study sites (FDR <0.001) among children breastfed at 3 months using Chi-square test as shown in the figure. (f) Odds ratio (95% CI) of perinatal factors and parental breastfeeding history for the presence of *B. infantis* in children breastfed at 3 months using logistic regression with adjustment of study site and sex. Colors represent the categories of these factors and only significant associations (p-value < 0.05) are solid.

Having identified *B. infantis* as a key species linked to SES-associated health outcomes, we next investigated the factors shaping its colonization and prevalence across populations. Although breastfeeding enriches *B. infantis*, it was detected in only 25% (N=352 out of 1387) ((Fig. 6c, d) of the CHILD Study cohort during the first year, mirroring findings from other North American cohorts that report reduced colonization compared to non-industrialized countries^42,43^. To explore factors influencing colonization beyond breastfeeding, we leveraged the geographical diversity of the CHILD Study, which includes samples from four Canadian cities. While 86% of infants were breastfed for at least three months, breastfeeding rates varied across sites (Extended Data Fig. 6a). Among these breastfed infants, *B. infantis* was the only *Bifidobacterium* species with significantly different prevalence across cities (FDR<0.001, Chi-square test), with Vancouver and Toronto showing the highest prevalence (37% and 30%) and Winnipeg and Edmonton the lowest (23% and 17%).These differences suggest that colonization is influenced by community-level factors beyond breastfeeding alone, possibly reflecting horizontal transmission influenced by the local microbiota meta-community^44^ (Figure 6e). Importantly, when we included Denmark in the analysis, we observed significant differences in the prevalence of several *Bifidobacterium* species (Extended Data Table 2). This may reflect technical differences in sampling age or broader population differences between the North American and European cohorts, such as environmental exposures, healthcare practices, or community microbiota composition^45,46^. Regardless, within the Canadian birth cohort, *B. infantis* remained the only *Bifidobacterium* species with variable prevalence, underscoring its distinct role in early colonization patterns.

Supporting this, both the abundance and prevalence of *B. infantis* were higher in breastfed infants during their first year than in those not breastfed (Fig. 6c, d). However, even after accounting for breastfeeding, *B. infantis* remained more abundant in children from higher-SES families in both the CHILD and COPSAC_2010_ cohorts (Extended Data Fig. 6b), suggesting additional influences on colonization. To further explore factors shaping *B. infantis* colonization, we used the breadth of perinatal and environmental data available from CHILD (Fig. 6f). Beyond environmental exposures, we were uniquely able to assess parental breastfeeding history, offering insights into potential cross-generational effects. Four factors were negatively associated with *B. infantis* prevalence: postnatal smoking, number of older siblings, cleaning product use, and carpet flooring. In contrast, neighborhood tree density, maternal asthma, and maternal—but not paternal— breastfeeding history were positively associated. These results suggest that horizontal transmission may be supported by both breastfeeding and community-level exposures, while maternal breastfeeding history points to an additional possible evolutionary mechanism favoring vertical transmission.

When considered in aggregate, these results suggest a protective role of *B. infantis* in child health and highlight the potential benefit for early-life supplementation, particularly in populations facing breastfeeding barriers. Beyond individual interventions, microbiota-targeted public health efforts such as antimicrobial stewardship, equitable access to safe green spaces, and smoke-free environments for infants should be prioritized to ensure all families benefit equally.

## Discussion

To effectively address environmental and lifestyle factors influencing disease, the scientific and medical communities must consider the compounded pressures of cultural, educational, and economic demands in industrialized societies^47^. This approach lays the foundation for reducing health disparities across all societal groups. In this study, we identify familial SES as a key driver of early-life exposures that shape microbiota composition and physiological development, setting the stage for long-term health trajectories. We defined SES using five measures, including income, educational attainment, and perceived social status, and found it was associated with more than half of the perinatal exposures infants in the CHILD study encountered, and associated with significant differences in the infant microbiota. This interplay was associated with health consequences ranging from asthma and excessive weight gain to behavioral challenges, illustrating how social and health inequities may emerge early in life.

The perinatal factors examined in this study encompassed parental health and diet, *in utero* exposures, early-life encounters during the first year, and conditions in both the home and broader environment. Each factor was linked to at least one health outcome at age five years, with many also associated with the infant gut microbiota as early as 3 months. Among these, breastfeeding emerged as the strongest associated factor, explaining much of the SES-associated differences in microbiota diversity and composition. Furthermore, breastfeeding may act as a buffer for the infant microbiota against environmental disruptions, and the ability to exclusively breastfeed an infant for the first 6 months of life, in accordance with current global health recommendations, may provide protection from the development of NCD-associated childhood risk factors in lower SES families. Breastfeeding is a highly conserved and adaptive behavior with extensive benefits for shaping both the microbiota and infant development, yet high-income countries have the lowest breastfeeding rates^18,39–41,48^. SES disparities in breastfeeding rates highlight modern challenges families face in maintaining breastfeeding, including pressure on parents to return to work quickly, and these challenges can be exacerbated by a lack of personal breastfeeding experience in previous generations^49–51^. Breastfeeding disparities therefore reflect a mismatch between societal and biological evolution, potentially contributing to the rise of NCDs in industrialized countries^7,52^. Our findings linking breastfeeding disparities to the rise of multiple NCDs highlight a disproportionate burden on families already experiencing social inequities and reaffirm arguments that the long-term health and economic benefits of breastfeeding are widely underestimated^53^. Addressing this issue requires societal-level policy and financial support, including generous paid parental leave, universal breastfeeding education and promotion in hospitals and birthing centers, and access to lactation specialists for ongoing support ^53,54^. Furthermore, when breastfeeding is not clinically possible, improving access to donor human milk or developing human milk-inspired alternatives is essential to preserve the benefits of a human milk diet for infants and their developing microbiota^55,56^.

Our analysis of the infant microbiota revealed that breastfeeding-associated changes were associated with protection against multiple adverse health outcomes at age five years. Notably, the species *B. infantis* was linked to reduced risk of atopy, asthma, and excessive weight gain. *B. infantis* has co-evolved with breastfeeding and is positively associated with several aspects of infant development^57–59^. However, it is present in only 25% of our cohort, a low prevalence echoed across North America, with some suggesting *B. infantis* loss may be a multi-generational consequence of industrialization^42,43^. Given the sensitivity of *B. infantis* to antibiotic exposure (Fig. 5a, Extended Data Fig. 3b), the combined rise in antibiotic use and formula feeding may have led to its reduction from our microbiota metacommunity over time, limiting its colonization even among breastfed infants. If this is the case, reducing barriers to a human milk diet in infancy should be complemented by the reintroduction of this important keystone species. While randomized controlled trials have already demonstrated some short-term benefits of supplementing *B. infantis* in breastfed neonates, the long-term impacts require further study and replicating stable *B. infantis* colonization and associated benefits in formula-fed infants remains a crucial step towards promoting equitable infant health^60–63^. Moreover, it is equally important to align public health efforts, such as antibiotic stewardship programs, that preserve *B. infantis* and other beneficial species within our communities. These efforts can help restore and maintain a resilient infant microbiota, supporting healthy development across society.

It is important to acknowledge the study’s strengths as well as its limitations, including the fact that the CHILD Study skews toward urban families with higher income and education. CHILD Study’s longitudinal, multi-center data includes comprehensive socioeconomic, perinatal, biological, and clinical data. This enabled us to associate a robust compound SES index with perinatal factors and multiple childhood NCDs to identify broadly protective factors. Stool samples collected at 3-month and 1-year visits also support a longitudinal, multi-analytic approach to link SES and perinatal factors with the infant gut microbiota using large-scale shotgun metagenomic sequencing. Our main analyses used a derived compound SES index, integrating several observed SES factors collected from questionnaires, which may have distinct effects. To ensure robust results, we performed sensitivity analyses using sub-cohorts and individual SES factors, exploring associations through various methods. What’s more, we replicated key findings in an independent cohort (COPSAC_2010_), including the associations between SES, breastfeeding, and multiple childhood NCDs, the mediating effect of breastfeeding on SES-associated infant gut microbiome, and the positive association between SES and *B. infantis*. While we found consistent effects of breastfeeding and SES on infant gut microbiota composition and individual taxonomy, we were limited to bacterial DNA profiling through shotgun sequencing. Moreover, we only profiled the gut microbiota, and we recognize the differences in the microbiota colonizing other body sites, including the lung and skin.

In conclusion, our findings demonstrate that familial SES significantly shapes the perinatal environment, influencing the infant gut microbiota and childhood health outcomes linked to NCD development. The protective association of SES was largely mediated by breastfeeding and the microbial species it supports, particularly *B. infantis*. Our results highlight the critical role of breastfeeding and beneficial microbes in fostering healthy childhood growth and well-being, while also emphasizing the social and economic pressures families face. These findings advocate for structural changes that prioritize evolutionarily conserved mother-infant interactions and ensure their accessibility to all families. Specifically, our study supports reducing barriers to breastfeeding, continued research into microbe-based therapies, and public health initiatives aimed at maintaining and restoring the infant microbiota in an equitable manner.

## Online Methods

### CHILD Study

#### Characteristics of the study population

The CHILD study is a prospective longitudinal birth cohort study, which enrolled 3,405 subjects since pregnancy from 4 largely urban study centers across Canada (Vancouver, Edmonton, Winnipeg, and Toronto) from 2008 to 2012^35^. Our study included 3,263 subjects eligible at birth (Extended Data Fig. 1), who were born at a minimum of 34 weeks of gestation and had no congenital abnormalities. CHILD Study followed children prospectively and collected detailed information on environmental exposures and clinical outcomes using a combination of questionnaires and in-person clinical assessments. Questionnaires related to environmental exposures, psychosocial stresses, nutrition and general health were administered at recruitment, prenatally, at 3, 6, 12, 18, 24, 30 months, and at 3, 4, and 5 years.

#### Socioeconomic status

In this study, we used the 2 continuous and 3 ordered categorical socioeconomic factors collected from questionnaires fulfilled by parents during pregnancy at 18 weeks and 1 year postnatal, which were the highest education of father and mother (4 ordinal levels: 1= complete high school, 2=complete college, 3= complete university, 4=master or PhD), annual household income (4 ordinal levels in Canadian Dollars: 1=0-49999, 2=50000-99999, 3=100000-149999, 4=over 150000), and continuous MacArthur scale of subjective social status score in Canada and community, where parents put themselves on the ladder, ranging from 1-10, of which 1 is the lowest and 10 is the highest. A total of 2,752 children had complete socioeconomic status data.

#### Early risk factors for multiple NCDs

In our study, adverse health outcomes collected at the 5-year visit were considered to be early risk factors for multiple NCDs, including overweight or obesity, physician diagnosis of asthma, atopic sensitization and emotional/behavioral problems.

##### Childhood overweight/obese

The weight and height measurements of children were performed by trained research assistants at clinic visits when children were 5 years old. Shoes and outerwear were removed for weight (Scaletronix scale) and height (standard stadiometer) measurements. BMI was calculated and age and sex BMI *z*-score (BMIz) was derived according to the World Health Organization (WHO) child growth standards^64^ for children younger than 5 years, and for 5–19 years for those children that were slightly older than 5 years old at the date of measurement. Children were classified as underweight with BMI z-scores less than -1, normal from -1 to 1, overweight from 1 to 2, and obese over 2^65^.

##### Childhood asthma

Childhood asthma was diagnosed (as Yes/Possible/No) by an expert study physician at the clinical assessment at the age of 5 years based on the CHILD Study’s published approach^35^. For this study, children were considered to have asthma only if the response was ‘Yes’ and the asthma phenotype was defined as comparing children with asthma (n=165) at 5 years versus children without asthma (n=2234) at 5 years, children diagnosed as “possible” (n=247) were excluded.

##### Childhood atopy

Children enrolled in the CHILD study were administered a skin prick testing (SPT) at the 5-year scheduled visit. Children were then diagnosed with IgE-mediated allergic sensitization (also referred to as atopy) based on SPT to multiple common foods and environmental inhalant allergens, using ≥2 mm average wheal size as indicating a positive test relative to the negative control. The allergens tested at 5-year visits include cat hair, the German cockroach, Alternaria tenuis, house dust mites, dog epithelium, cow’s milk, peanut, egg white, soybean, *Cladosporium*, *Penicillium*, *Aspergillus fumigatus*, trees, grass, weeds, and ragweed. Glycerin and histamine served as the negative and positive controls, respectively^26^.

##### Childhood emotional/behavioral problems

Childhood emotional/behavioral problems is defined based on Children Child Behavior Checklist (CBCL)^66^, which is a widely used and validated questionnaire to assess behavioral and emotional problems. Parents report on the frequency of child problems or behaviours over the last 6 months using a 3-point Likert scale. Higher scores indicate greater symptoms. In this study, children with CBCL T-score > 60, the borderline elevation, in either externalizing or internalizing problems at 5 years of age were defined as children with emotional/behavioral problems.

#### Perinatal factors and exposomes

##### Pregnancy

Relevant exposures during pregnancy collected by the CHILD study included gestational diabetes during pregnancy, maternal distress during pregnancy (reported perceived distress (stress or depressive symptoms) using the 10-item PSS)^67^, maternal antibiotic during pregnancy and prenatal smoking (maternal and second-hand). These variables were derived from questionnaires during pregnancy at 18 weeks.

##### Birth

Relevant exposures during delivery collected by the CHILD study included the mode of delivery (vaginal versus c-section), the season of birth (spring, summer, fall and winter), intrapartum antibiotic exposure, gestational age and gestational age-adjusted birth weight z-score^68^, derived from birth charts or questionnaires. Child sex was also collected from birth charts.

##### Postnatal

Postnatal exposures in this study included any breastfeeding duration, exclusive breastfeeding status at 6 months, regular childcare attendance at 3 months and 1 year (indicated by regularly going to a location at least 1 hour per day on average or at least 7 hours total in a week), the number of older siblings, child antibiotic use in the first year of life, cat and dog ownership and postnatal smoking (maternal and second-hand). These variables were derived from postnatal standardized questionnaires. In analyses of infant gut microbiota, child antibiotic use was defined as antibiotic exposure before the stool sample collection age and breastfeeding was defined as children were still breastfed at the stool sample collection age.

##### Home Environment

Home environmental factors included home type (multiple-family or single-family), and the medium or high mould, carpet floor, and having potted plant, which were summarized from kitchen, bathroom, kid room, mother room and living room, clutter home (cluttered furniture or decoration generally at home), and frequency of use score of cleaning products. These variables were summarized from questionnaires and notes during the home assessment by the research assistant when the child was 3–4 months of age. The frequency of use score of cleaning products (FUS) was derived by summing the scores assigned to their questionnaire responses for the 26 categories of products in the previous CHILD study^69^.

##### Neighborhood Environment

Physical Environmental factors included individual exposures to nitrogen dioxide (NO_2_), dwelling density z−score, tree density and urbanicity (rural vs. urban). NO_2_ is a measure of traffic-related air pollution, estimated from city-specific land use regression models^70^ across pregnancy and in the first year of life. Dwelling density z-scores were calculated based on the number of dwellings per hectare using data from the CANUE urban environmental health datasets during pregnancy at 18 weeks. Urbanicity was determined by the rural/urban location at the home address where the family resided the longest around the time of birth (period covers pregnancy and first year of life) based on 20006 census data^71^. Tree density was defined using public tree census data. Tree locations were geocoded and averaged within a 250m buffer centered on the study participants’ home addresses during pregnancy.

##### Parental Diet and Health

Parental diet factors included maternal ultra-processed food intake. We used the NOVA classification system to define maternal ultra-processed food intake based on the extent and degree of food processing^72^. Parental health factors included maternal and paternal asthma and atopy, maternal BMI, which were derived from questionnaires during pregnancy at 18 weeks or 1 year postnatal, and maternal distress (reported perceived distress) at 1 year postnatal.

##### Genetic ancestry

Genetic ancestry was represented by the first 3 PC variables associated with self-reported parental ancestry derived from genotyped cord blood data using the principal component analysis^73^.

#### Stool Sample Collection

A subsample of 1,479 children had shotgun metagenomic data processed from fecal samples collected at 3 months (n=1,422), 12 months(n=1,426), or 3 and 12 months of age (n=1,369) (Extended Data Fig. 1). Sample collection and sequencing were performed as previously described^24^. Briefly, stool samples from diapers were collected at a home visit at around 3 months [mean (SD), 3.8 (1.1) months] and a clinic visit at around 1 year [mean (SD), 12.4 (1.3) months]. Samples were briefly stored at 4 °C and then aliquoted into four 2-mL cryovials using a stainless steel depyrogenated spatula and were frozen at −80 °C. CHILD recorded the time between stool collection and long-term storage. This processing time and the age of children at the time of stool sample collection were adjusted for in our statistical analysis. Samples collected for children over 1.5 years of age or samples with processing time higher than 100 hours were excluded.

#### Gut Microbiota Characterization and Pre-processing

Shotgun metagenomic sequencing data with an average depth of 5 million reads per sample was generated by Diversigen (Minneapolis, MN, USA) from fecal samples. DNA was extracted from faecal samples using the MO Bio PowerSoil Pro with bead beating in 0.1mm glass bead plates. High-quality input DNA was verified using Quant-iT Picogreen. Libraries were then prepared and sequenced on an Illumina NextSeq using single-end 1 x 150 reads. Low quality (Q-Score<30) and length (<50) sequences were removed, and adapter sequences trimmed. Host and low-quality reads were removed, and only samples with at least 1 million remaining reads or more were retained for downstream analysis. Shotgun metagenomic reads were mapped using the bioBakery 3 pipeline^74^ to identify taxonomic (species and strain level) and functional features within each sample. To generate the abundance of Bifidobacterium longum subspecies, MetaPhlAn database was customized with a set of maker genes including 119 *Bifidobacterium logum. subspecies infantis* and 128 *Bifidobacterium longum subspecies longum* markers using MetaPhlAn instructions (https://github.com/biobakery/MetaPhlAn/wiki/MetaPhlAn-4) and then MetaPhlAn4 was used with the --index and --bowtie2db parameters and the customized marker-gene database^75^ and HUMAnN 3 was used for functional profiling.

### COPSAC_2010_ cohort

#### Characteristics of the study population

Similar to the CHILD study, the COPSAC_2010_ cohort is an independent population-based mother-child cohort of 700 children and their families in Denmark. Families were enrolled in pregnancy and children were followed prospectively by COPSAC_2010_ study physicians. All biosamples were collected by nurses and clinical diagnoses were assessed during clinical visits at 1 week, 1, 3, 6, 12, 18, 24, 30, and 36 months, yearly until the age of 6 and again at age 8 and 10 years. Information on duration of exclusive and total breastfeeding period was recorded during the scheduled visits to the research clinic^76,77^.

#### Socioeconomic status

The COPSAC_2010_ cohort collected three measures of socioeconomic at1 week after birth, including household income covering last 3 months of pregnancy (4 ordinal levels in Danish Krone: 1=0-150,000, 2=150,000-200,000, 3=200,000-250,000, 4=over 250,000) and the highest education of mother and father mother (4 ordinal levels: 1=complete high school or college, 2=complete tradesman, 3=complete medium academic, 4=complete university).

#### Early risk factors for multiple NCDs

In the COPSAC_2010_ cohort, information on physician-diagnosed asthma, duration of exclusive and total breastfeeding period was obtained during the scheduled visits to the research clinic. The blood sample and Strengths and Difficulties Questionnaire (SDQ) was assessed at 6-year clinic visit.

##### Childhood overweight/obese

Weight was measured by calibrated digital weight scales and without clothes. Length was measured until the age of 2 years using an infantometer (Kiddimeter; Raven Equipment Ltd, Dunmow, Essex, England). After the age of 2 years, height was measured using a stadiometer (Harpenden, Holtain Ltd, Crymych, Dyfed, Wales) which was calibrated yearly. BMI was calculated as weight (kg)/length^2^ (m^2^) and converted into age-and-sex-standardized z-scores using the R package ‘zscorer’^78^. Same as CHILD cohort, children were classified as underweight with BMI z-scores less than -1, normal from -1 to 1, overweight from 1 to 2, and obese over 2^65^ at age 5 years.

##### Childhood asthma

Asthma was diagnosed on the basis of a previously detailed quantitative symptom algorithm requiring all of the following criteria^79–81^: (i) verified diary recordings of five episodes of troublesome lung symptoms within 6 months, each lasting at least three consecutive days; (ii) symptoms typical of asthma including exercise-induced symptoms, prolonged nocturnal cough, and persistent cough outside of common colds; (iii) need for intermittent rescue use of inhaled β2-agonist; and (iv) response to a 3-month course of inhaled corticosteroids and relapse upon ending treatment^79^. Remission of asthma was defined by 12 months without relapse upon cessation of inhaled corticosteroid treatment. For analyses, we used the ever diagnosed asthma at age 5 years.

##### Childhood atopy

Specific IgE levels were determined at ages 0.5, 1.5, and 6 years byusing a screening method (ImmunoCAP, Phadiatop Infant™, ThermoFisher Scientific)^76^. The 6-month blood sample was further analysed using ImmunoCAP ISAC® measuring 112 components from 51 different allergen sources. Levels ≥ 0.3 ISAC Standardized Units(ISU) were considered indicative of allergic sensitization. The 6-year blood sample was analysed for dog, cat, grass, birch, mugwort, D pteronyssinus, moulds, egg, milk,wheat flour, and peanut by ImmunoCAP, using any level ≥0.35kUA/L as indicative of sensitization, that is elevated specific IgE^82^.

##### Childhood emotional/behavioral problems

Childhood emotional/behavioral problems is defined based on SDQ at 6 years of age, which is a widely used screener for detecting mental health difficulties^83^. Higher scores indicate greater symptoms. To be comparable to externalizing (Conduct and Hyperactivity) or internalizing (Emotional and Peer problems) problems based on CBCL assessment in CHILD cohort. Children with score higher than clinical cut-off scores for the subscales (out of a possible 10)^83^: Emotion ≥ 5, Conduct ≥ 4, Hyperactivity ≥ 7, Peer Problems ≥ 4, were defined as children with emotional/behavioral problems.

#### Stool Sample Collection

Fecal samples were collected either at the research clinic or by the parents at home using detailed instructions. Each sample arrived at the laboratory was mixed on arrival with 1 ml of 10% (v/v) glycerol broth (SSI, Copenhagen, Denmark) and frozen at −80°C. DNA was extracted using the PowerMag Soil DNA Isolation Kit (Qiagen) and NucleoSpin Stool Kit (Macherey-Nagel). No differences were found between the different DNA extraction kits after comparison. Before library preparation, the DNA was quantified by Tecan Infinite F Nano+ Plate Reader using Quant-iT dsDNA BR Assay Kit. The enzymatic fragmentation of DNA and library construction was conducted by Tecan DreamPrep NGS using Celero EZ DNA-seq Core Module Kit. The fragmented DNA was amplified using polymerase chain reaction (PCR). Short and large DNA fragments were removed using double-sided magnetic bead size selection (AMPure XP, Beckman Coulter). Adapter sequences from Celero 96-Plex Adaptor Plate were added to each sample during library construction. The final concentration for each library was quantified by Tecan Infinite F Nano+ Plate Reader using NuQuant NGS Library Quantification Module and Qubit. The final fragment distribution is evaluated using a Fragment Analyzer 5200 (Agilent). Qubit and TapeStation were used to determine the concentration of the final library before sequencing. The library was sequenced using 2 × 150 bp paired-end sequencing on an Illumina NovaSeq 6000 platform (Illumina, San Diego, CA, USA). The shipment time and the age of children at the stool sample collection time were adjusted for in our statistical analysis. Similar to the CHILD study, samples collected from children over 1.5 years of age or those with shipment times exceeding 100 hours were excluded.

#### Gut Microbiota Characterization and Pre-processing

Quality control of raw FASTQ files was performed using KneadData (v. 0.6.1) to remove low-quality bases and reads derived from the host genome as follows: Using Trimmomatic (v. 0.36), the reads were quality trimmed by removing Nextera adapters, leading and trailing bases with a Phred score below 20, and trailing bases in which the Phred score over a window of size 4 drops below 20. Trimmed reads shorter than 100 bases were discarded as low-quality reads. Reads that mapped to the human reference genome GRCh38 (with Bowtie2 v. 0.2.3.2 using default settings)^84^ were also discarded. Read pairs in which both reads passed filtering were retained; these were classified as high-quality non-host (HQNH) reads. Shotgun metagenomic reads were mapped using the bioBakery 3 pipeline^74^ to identify taxonomic (species and strain level) and functional features within each sample. Similar to the CHILD cohort, the MetaPhlAn database was customized to generate the abundance of *Bifidobacterium longum subspecies*^75^.

### Statistical Analysis

Data analysis was conducted in R (version 4.3.1). We summarized the SES latent factor using confirmatory factor analysis (CFA) using SES observed factors, including household income, the highest education of mother and father, and the MacArthur scale of subjective social status in Canada, and the community collected during pregnancy at 18 weeks. The COPSAC_2010_ SES latent factor was generated using SES observed factors, including household income, the highest education of mother and father. Models were estimated using “cfa” function included in the “lavaan” R package^85^. Household income and the highest education of mother and father were treated as ordered categorical variables in the models. Models were stepwise updated using the function “modindices”^85^ which identified significant correlations between two covariant variables (chi-square p-value < 0.05) to improve the model fit. “lavPredict” function was used to compute the estimated values (’factor scores’) for the SES latent variable in the model, which was used to represent summarized SES index for later analysis. Item cluster analysis was performed to confirm the associations across the observed SES factors using “iclust” function from “psych” R package^86^ to cluster SES factors and perinatal factors and exposomes using correlations with pairwise deletion of samples. Least square regression models with adjustment of study center and sex were applied to explore the associations between SES, childhood NCDs and each perinatal factor. The confidence interval was estimated using the “confint” function from the “stats” R package. Factors with p-values < 0.05 were defined as significant. P-values were without adjustment of multiple comparisons, while q-values (FDR) represent p-values with adjustment of multiple comparisons using Benjamini-Hochberg (BH) method. Ethnicity or genetic ancestry is well-known to be associated with SES. To fully understand the effects of SES and explore the robustness of the summarized SES index, we performed sensitivity analyses with and without adjustment of the genetic ancestry using the first 3 genetic ancestry PCs. In addition, sensitivity analyses using 1-year summarized SES index, each study center, and each of the observed 5 SES factors were also performed, in which the ordinal SES categorical variables were treated as numeric variables based on their orders.

Shotgun metagenomic data was rarefied and microbial community structure was measured by using the within-individual species diversity (α-diversity) by Shannon index. To test the associations between infant gut microbiota and SES index and perinatal factors, Euclidean distance (“vegan”) was calculated on the modified centered log-ratio (mCLR)-transformed^87^ relative abundance of species. mCLR transform applies central log transformation to non-zero values and adds a pseudocount of 0.1 and the minimal value to them, and the previous exact zero relative abundance entries in the OTU table remained zero. Then, a marginal Permutational Multivariate Analysis of Variance (PERMANOVA) was performed using “adonis2” function from “vegan” R package across 1000 permutations with adjustment of stool sample collection age, processing time and a strata of study center^88^. The beta diversity based on the Euclidean metric was used to perform principal coordinates analysis on mCLR-transformed relative abundance of species. The association between the first three principal coordinates (PCoAs) and SES index were tested using Wilcoxon tests (compare PCoAs in children with the second, third, and fourth quartiles of familial SES index to children with the first quartile of SES index) and linear mixed effect models (test association between PCoAs and continuous SES index) with study center as a random effect and adjustment of stool sample collection age and processing time. Statistical significance was defined as a p-value < 0.05. The top and/or SES-associated PCoAs were graphed to visualize the sample group relationships, where the arrows represent the strength and direction of the top 10 factors that explain microbiome variation using Pearson correlation. As we hypothesized that the effect of SES on overall infant gut microbiota would be through perinatal factors, structural equation modelling (SEM), a statistical technique used to evaluate how well observed data match hypothesized causal relationships, was then used to assess the potential mediations using the R package “lavaan”^85^. For model specification, we simultaneously estimated the mediation effects (indirect effects) of perinatal factors on SES-associated infant gut microbiota composition using PCoA at 3 months and 1 year, respectively (PCoA1 for 3-month and PCoA3 for 1-year infant microbiota). Using COPSAC_2010_ cohort, the mediation effect of breastfeeding was estimated on SES-associated infant gut microbiota composition using PCoA3 at 1 month and 1year, respectively. All regressions in SEM models were adjusted for study center and regressions with microbiome data were also adjusted for stool sample collection age and processing time. In regression models and SEM models, all categorical variables were replaced by dummy variables, i.e. study center was replaced by Vancouver, Toronto and Edmonton. All univariable and SEM models, which focused on individual perinatal factors, excluded observations without pairwise data (considered missing at random). To explore differences in perinatal factors’ effect on infant gut microbiota between children with and without breastfeeding, we compared the beta dispersion and repeated our PERMANOVA analysis on samples stratified by breastfeeding status. The breastfeeding (BF) group included children who were not breastfed at the stool sample collection age and no breastfeeding (No BF) group included children who were either never or not breastfed at the 3-month visit. To account for differences in sample size, a bootstrapping test (randomly selecting the same number of samples as the smaller group with replacement for 100 runs) was applied. Factors with p-value based on PERMANOVA analysis less than 0.05 for at least 80% of runs were considered significant. Moreover, to explore the effect of breastfeeding on childhood NCDs for children with different familial SES, a compound variable was generated based on the lower or upper half of the SES index and exclusive breastfeeding status to 6 months. Then, logistic regression with adjustment of study center and sex was applied to examine its association with the risk of at least one SES-associated childhood risk factors for NCDs (childhood overweight/obesity, asthma and emotional/behavioral problems at 5 years of age) with children from lower half SES index family and not breastfed to 6 months as a reference group. Statistical significance was defined as a p-value < 0.05.

The “Phyloseq” package^89^ was used to preprocess the metagenomic taxonomy table. To identify bacterial species that were significantly associated with SES, perinatal factors, and childhood NCDs, linear mixed-effect models (MaAsLin2) models were applied to the mCLR transformed relative abundance of each species and Metacyc pathways. Only species and Metacyc pathways detected in at least 10% of samples were used. Longitudinal models were performed with and without interaction between time and factor, with the study center and subject ID as random effects. For each species and factor, the formula of model with interaction is Species (mCLR-transformed relative abundance) ∼ β_a1_*factor + β_a2_*stool sample collection age + β_a3_*SES*stool sample collection age+ β_a4_*processing time/shipment time + 1|(site+subject)), in which β_a3_ is for the slope effect, measuring the change of factor over time, while β_a1_ is for the baseline effect, measuring the baseline difference. The formula of the model without interaction is Species ∼ β_b1_*factor + β_b2_*stool sample collection age+ β_b3_*processing time/shipment time + 1|(site+subject), in which β_b1_ measures the effect of the factor on overall colonization of the species. All MaAsLin2 models were adjusted for stool sample of collection age and processing time/shipment time. Benjamini-Hochberg procedure was used to correct p-values, and tests with MaAslin2 qval (FDR) < 0.1 were considered to be significant. Then, to test the mediation effect of infant microbiome on SES-associated child health outcomes, again SEM model was applied, in which we conceptualized the infant gut microbiome (i.e. latent variable) using CFA, including the 30 SES-associated species at 3 months and 1 year. “modindices” was used to identify significant (chi-square p-value < 0.05) correlations across species to improve the model fit. All regressions in SEM models were adjusted for study center and sex, and regressions with microbiome data were also adjusted for stool sample collection age and processing time/shipment time. Models with CFI exceeding 0.9, RMSEA lower than 0.05, and SRMR lower than 0.08 are considered a good fit^90^.

## Acknowledgments

We are grateful to all the families who took part in this study and the whole CHILD team, which includes interviewers, nurses, computer and laboratory technicians, clerical workers, research scientists, volunteers, managers, and receptionists. We are grateful to the children and families who participated in the COPSAC2010 cohort study for their dedication and support. Furthermore, we acknowledge and appreciate the unique efforts of all members of the COPSAC research team.

## Funding

S.E.T. holds a Tier 1 Canada Research Chair in Pediatric Precision Health and the Aubrey J. Tingle Professor of Pediatric Immunology.

D.L.Y.D. is funded by a Canadian Institute of Health Research Frederick Banting and Charles Best Canada Graduate Scholarship Doctoral Award (CIHR CGS-D) and the University of British Columbia Four Year Doctoral Fellowship (4YF).

M.B.A. holds a Tier 2 Canada Research Chair in Early Nutrition and the Developmental Origins of Health and Disease and is a Fellow if the CIFAR Humans and the Microbiome Program.

P.S. holds a Tier 1 Canada Research Chair in Pediatric Asthma and Lung Health.

P.J.M holds Women’s and Children’s Health Research Institute.

Funding for this specific study came from Genome Canada and Genome British Columbia (grant to S.E.T. [274CHI]) with additional support BC Children’s Hospital Research Institute and Foundation, as well as the Provincial Health Services Authority.

COPSAC is supported by a variety of private and public research funds, listed on www.copsac.com.

## Author contributions

Conceptualization: D.L.Y.D, C.P, and S.E.T.

Methodology: D.L.Y.D and C.P.

Investigation: D.L.Y.D.

Visualization: D.L.Y.D and C.P.

Funding acquisition: S.E.T., P.S, P.J.M, M.B.A., T.J.M, E.S.

Project administration: S.E.T. and P.S.

Supervision: C.P and S.E.T.

Writing – original draft: D.L.Y.D, C.P and S.E.T.

Writing – review & editing: D.L.Y.D, M.B.M, C.H, H.S, K.M, S.C.C, D.J.K, Q.D, T.J.M, P.J.M, B.B.F, E.S, H.L., D.M.P, P.S, M.B.A, C.P, and S.E.T.

### Data availability

The accession number for the shotgun metagenomic data reported in this paper is BioProject accession (NCBI): PRJNA838575. Code for analyses and figures are provided in the supplement. The informed consent obtained from the CHILD participants, in addition to the CHILD Inter-Institutional Agreement (IIA) which has been executed between the five Canadian institutions responsible for the study, govern the sharing of CHILD data. Data described in the manuscript are available by registration to the CHILD database (https://childstudy.ca/childdb/) and the submission of a formal request. All reasonable requests will be accommodated. More information about data access for the CHILD Cohort Study can be found at https://childstudy.ca/for-researchers/data-access/. Researchers interested in collaborating on a project and accessing CHILD Cohort Study data should contact. COPSAC sequencing data is available in the Sequence Read Archive (SRA) under accession no. PRJNA715601. Individual-level data is protected under Danish and European law that prohibits publication even in pseudonymized form. However, data can be made available to researchers under a data processing agreement by contacting COPSAC’s Data Protection Officer.

### Code availability

The code for the study is provided in the Supplementary Files.

