## Supplementary material for "Breastfeeding limits the adverse impact of socioeconomic status on child health by modifying the infant gut microbiome"

#### Extended Data

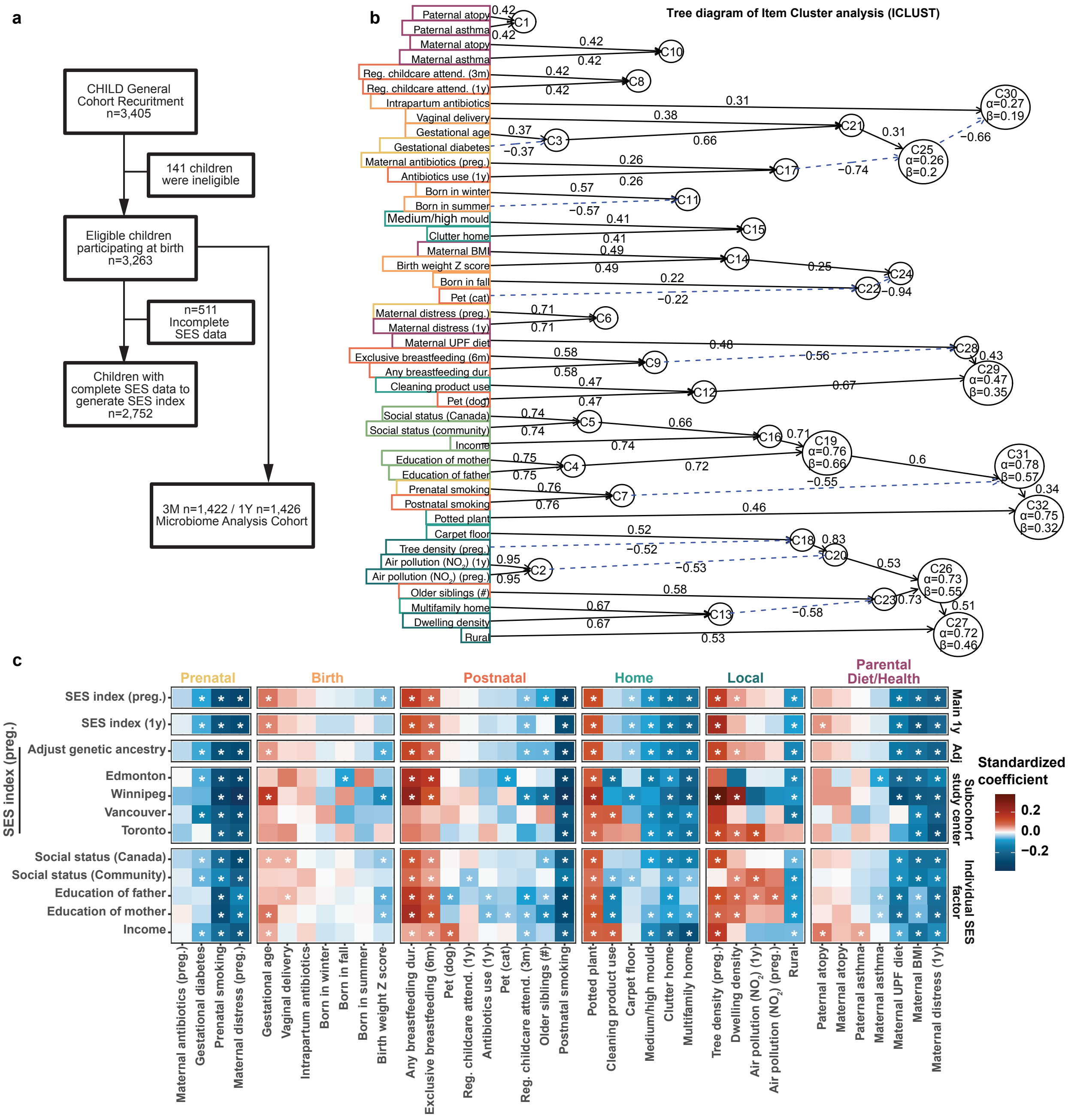


**Extended Data Fig. 1. Associations across SES, perinatal factors, and childhood health outcomes.** (a) Consort diagram of CHILD Cohort. (b) Tree diagram of the item cluster analysis (ICLUST) to cluster observed SES factors and perinatal factors and exposomes based on their Pearson correlations. Cluster 19 (C19) was found to include the 5 observed SES factors with Cronbach's alpha values >0.7, indicating internal consistency within the calculated clusters. (c)

Heatmap of the associations between summarized SES index at enrollment and perinatal factors, and sensitivity analysis using summarized SES index based on observed SES factors collected at 1 year, with adjustment of genetic ancestry, sub-cohort analyses for each study center and individual observed SES factors using regression models.

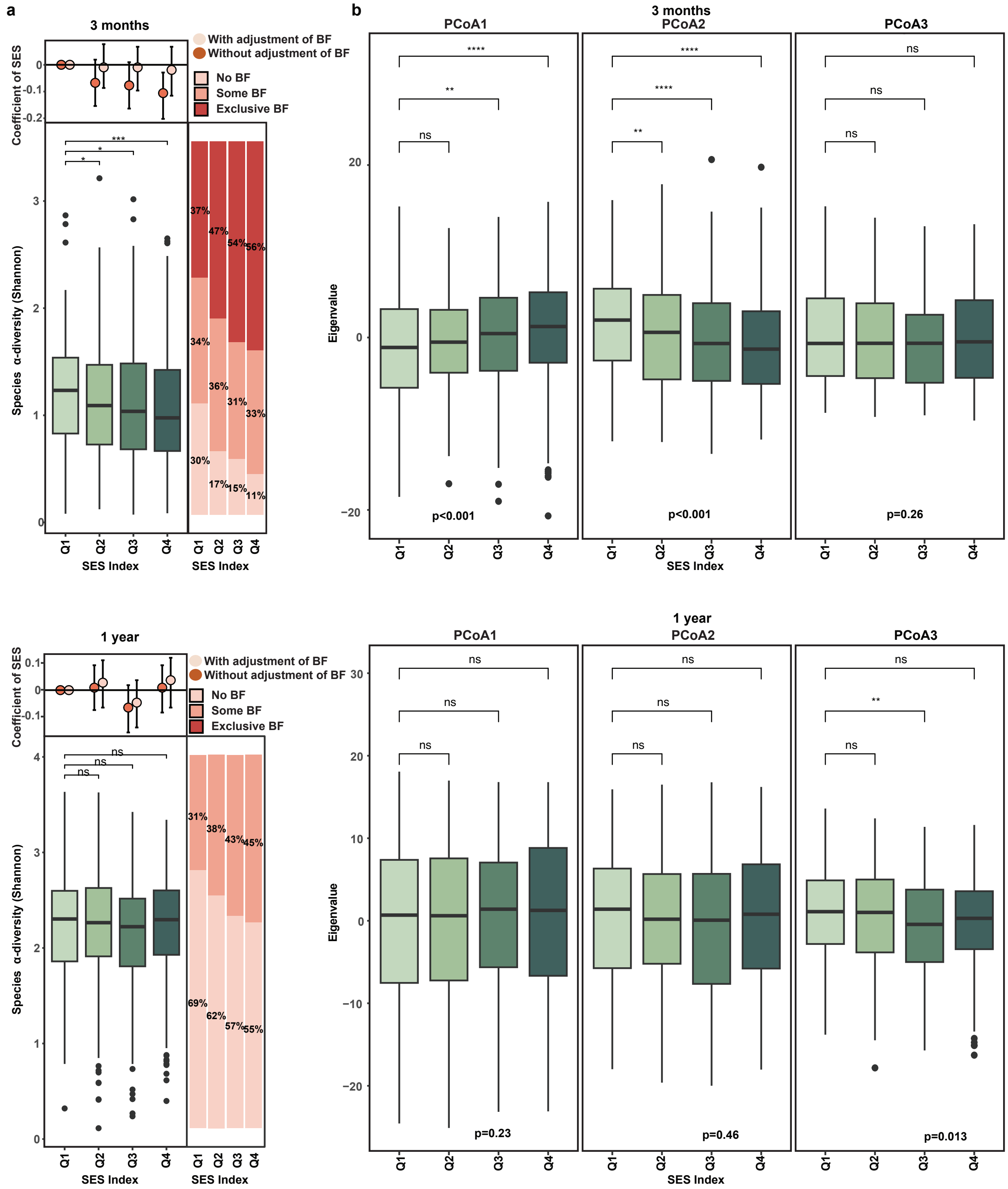


**Extended Data Fig. 2. SES is associated with infant microbiome composition at 3 month and 1 year.** (a) Differences in gut microbiota species α-diversity (Shannon index) at 3 months and 1 year across children with different quartiles of SES index. SES Q1 includes samples with the lowest 25% SES index and SES Q4 includes samples with the top 25% SES index. The right-side stacked bar plots showed the proportion of some (dark pink) and exclusive (orange) breastfeeding across SES quartiles. The top panels show the associations between species α-diversity and SES quartiles using linear regression models with (pink) and without (orange) adjustment of breastfeeding. Linear regression models were all adjusted for stool sample collection age and processing time and with study site as a random effect. (b) Boxplot of the top 3 PCoAs across SES quartiles. Differences across SES quartiles compared to the first quartile of SES were estimated based on Wilcoxon test. ns $p>0.05$, * $p<0.05$, ** $p<0.01$, and *** $p<0.001$. The box displays 25th, 50th (median), and 75th percentiles, with whiskers extending 1.5 * IQR. P-values at the bottom of each figure were based on linear regression models on continuous SES index with study site as a random effect and adjustment of stool sample collection age and processing time.

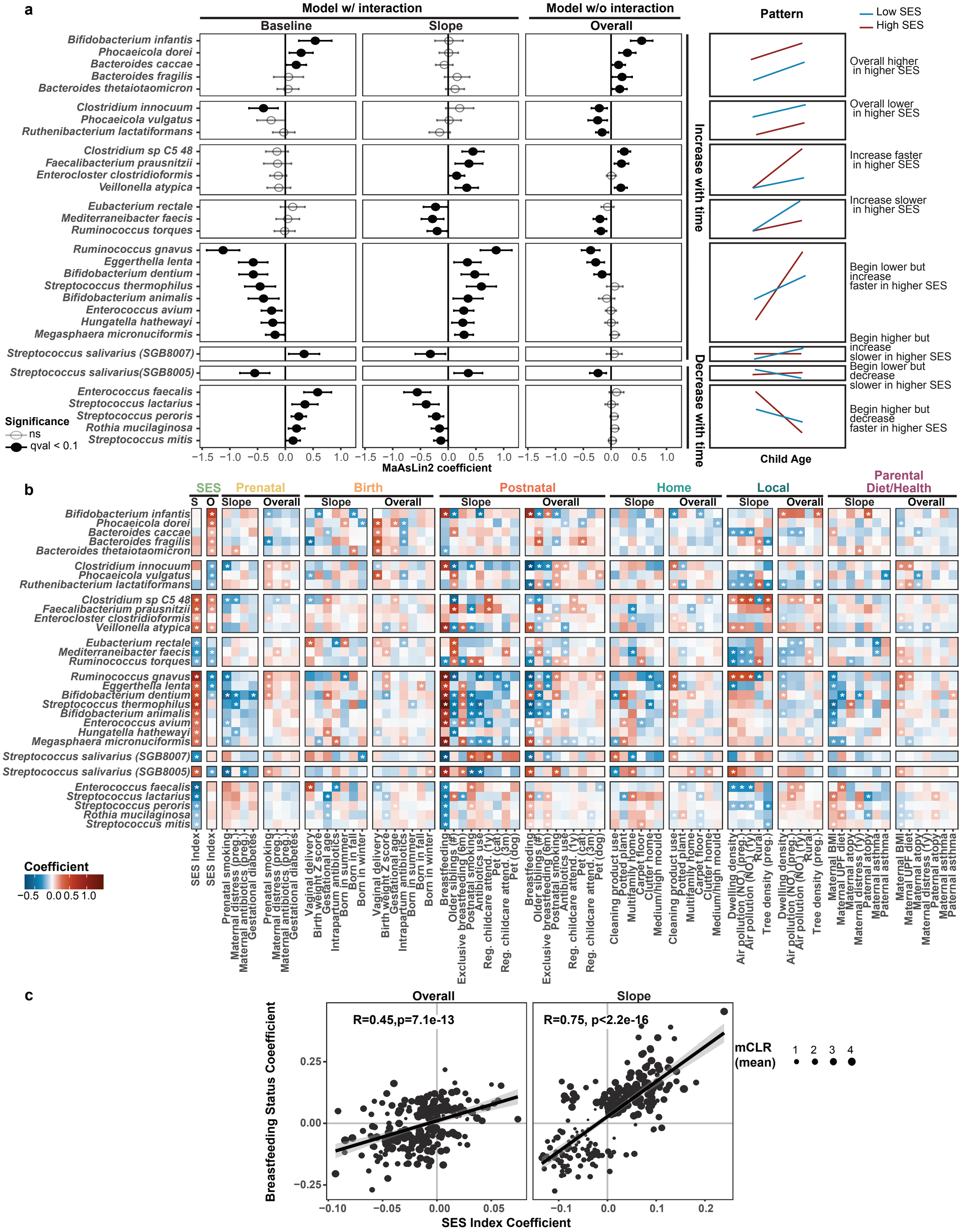


**Extended Data Fig. 3. Species and Metacyc pathways associated with SES, breastfeeding, and perinatal factors and exposomes.** (a) Baseline and slope effect (model with interaction) and overall effect (model without interaction) of SES index for species significantly associated with

SES (FDR of slope effect or overall effect < 0.1) across 3 months to 1 year based on MaAslin2 models. Data were presented as standardized coefficients of SES ± 1.96*standard error of the mean. The right-side panel summarizes the pattern of SES and time effect on each species across child age, in which the red line represents higher SES and the blue line represents lower SES and there is an interaction if the slope effect is significant. (b) Heatmap of the standardized slope and overall effect of perinatal factors on SES-associated species based on MaAslin2 models (FDR <0.1). Red represents positive association, while blue represents negative association. Stars represent significant associations (FDR < 0.1). (c) Correlation plot of overall effect (MaAslin2 models without interaction measuring overall colonization) and slope effect (MaAslin2 models with interaction measuring change over time) between SES and breastfeeding for Metacyc pathways significantly associated with either SES or breastfeeding (FDR of slope or overall effect < 0.1). Dot size represents the mean of relative abundance of Metacyc pathways across 3 months and 1 year. All MaAslin2 models used subject ID and study center as random effects and were adjusted for stool sample collection age and processing time.

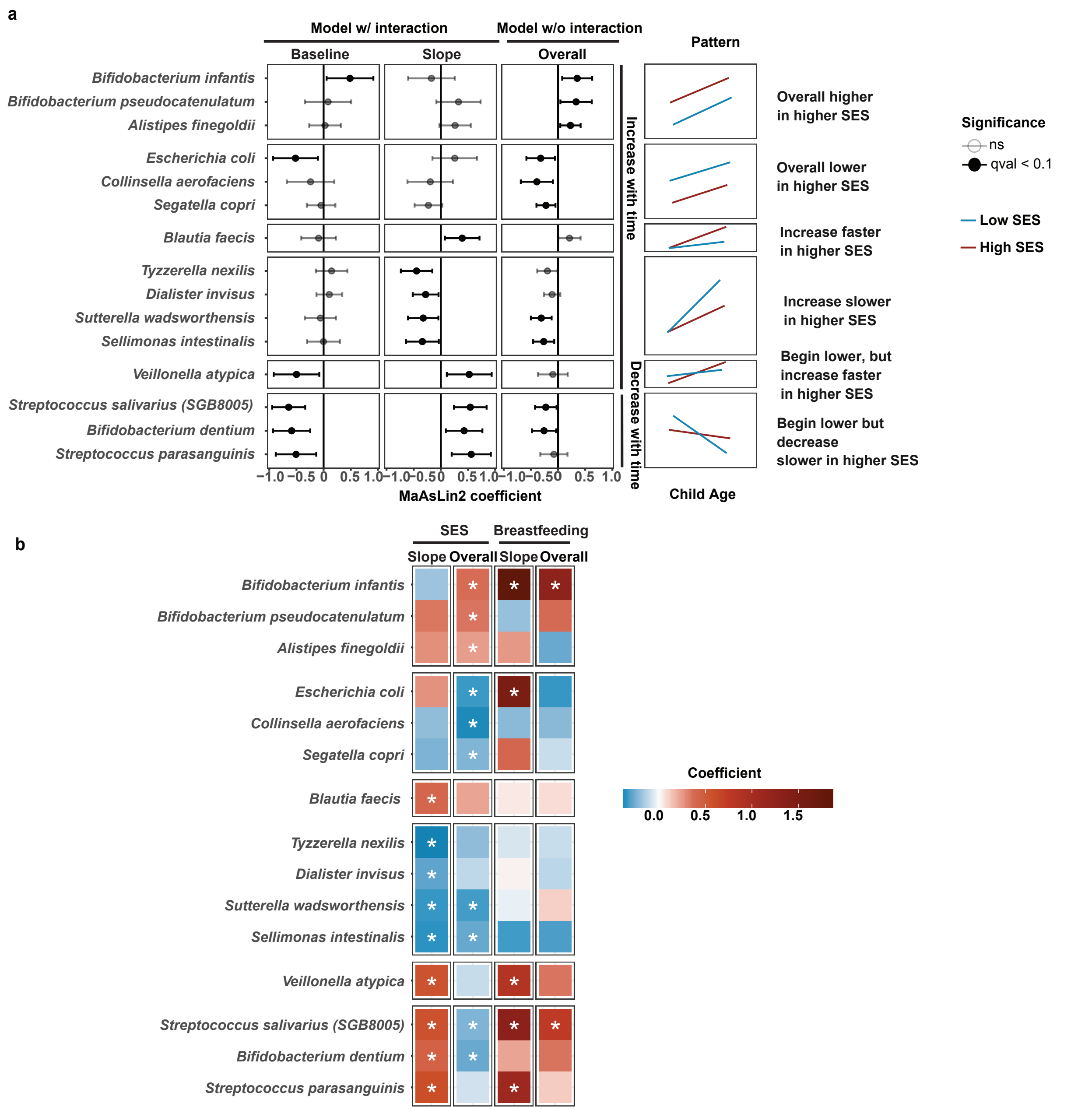


**Extended Data Fig. 4. Species significantly associated with SES and breastfeeding in COPSAC$_{2010}$ cohort.** (a) Baseline and slope effect (model with interaction) and overall effect (model without interaction) of SES index for species significantly associated with COPSAC$_{2010}$ SES index (FDR of slope effect or overall effect < 0.1) across 1 month to 1 year based on MaAslin2 models. Data were presented as standardized coefficients of SES ± 1.96*standard error of the mean. The right-side panel summarizes the pattern of SES and time effect on each species across child age, in which the red line represents higher SES and the blue line represents lower SES and there is an interaction if the slope effect is significant. (b) Heatmap of the standardized slope and overall effect of SES and breastfeeding on SES-associated species based on MaAslin2 models (FDR <0.1). Red represents positive association, while blue represents negative association. Stars represent significant associations (FDR < 0.1).

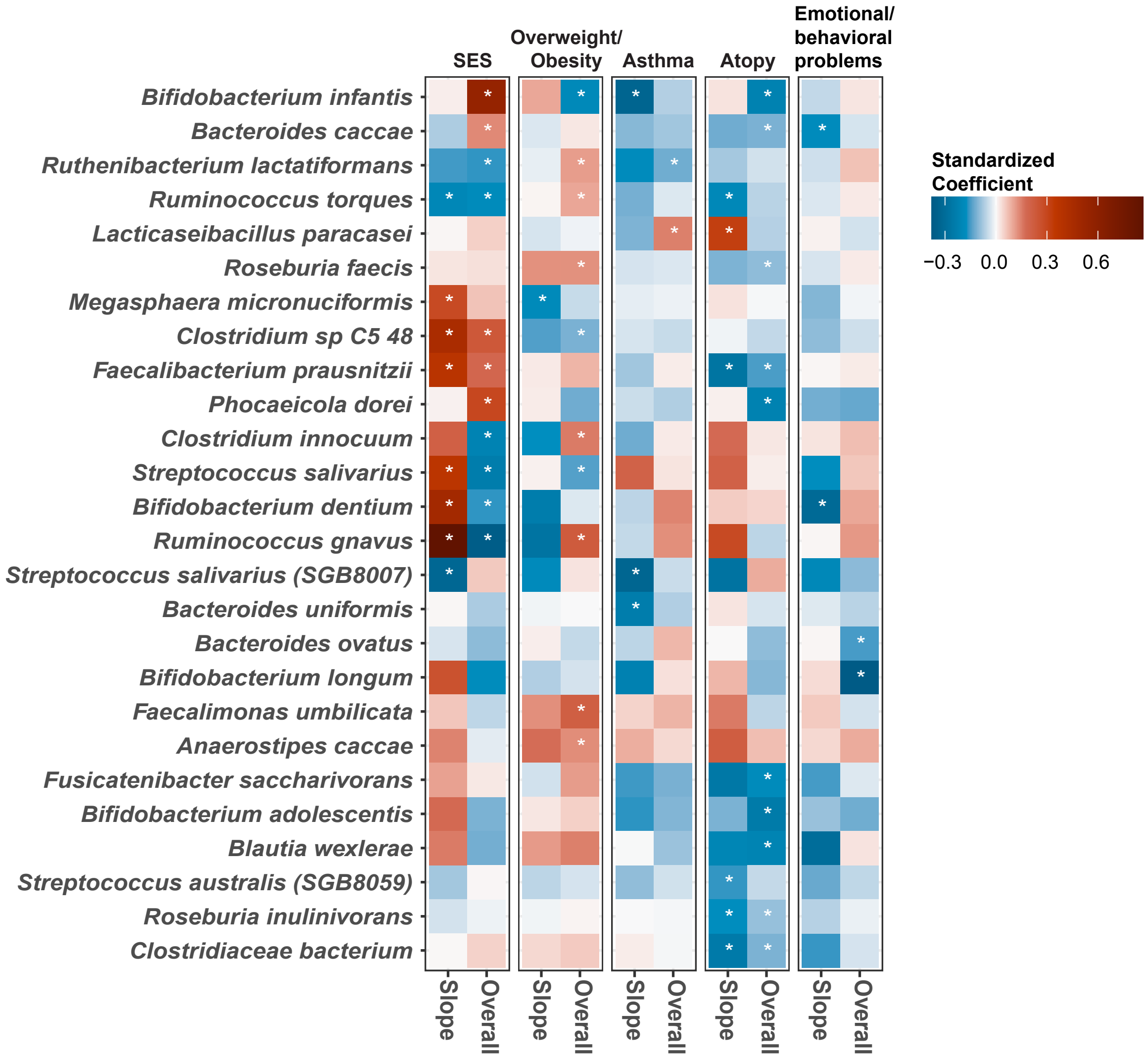


**Extended Data Fig. 5. Heatmap of standardized coefficient of SES index and childhood NCDs for species significantly associated with at least one of four childhood NCDs (FDR < 0.1) using MaAslin2 models with (slope effect) and without (overall effect) interaction.** All MaAslin2 models used subject ID and study center as random effects and were adjusted for stool sample collection age and processing time. Red represents positive association, while blue represents negative association. Stars represent FDR < 0.1.

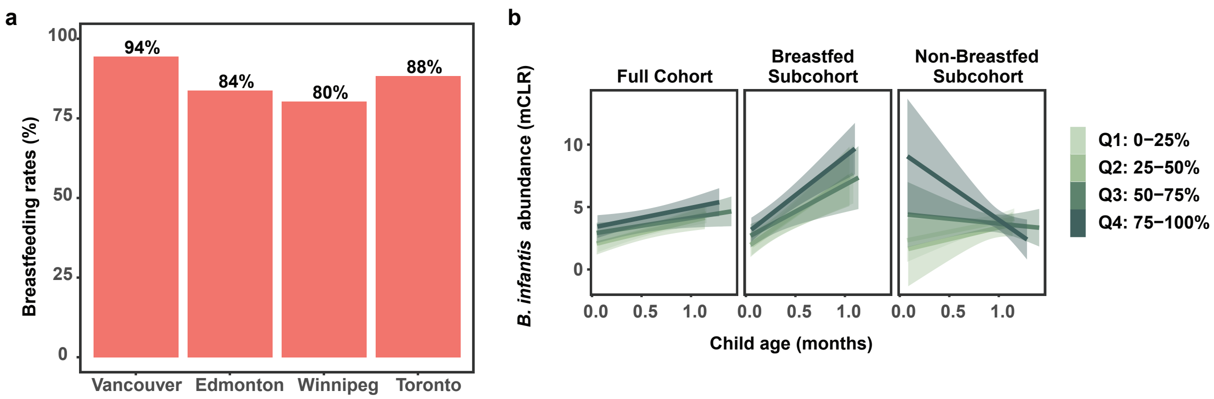


**Extended Data Fig 6. Breastfeeding rate in CHILD cohort and *B. infantis* abundance in COPSAC$_{2010}$ cohort.** (a) Breastfeeding rate (some breastfeeding at least 3 months) across CHILD study sites. (b) Relative abundance of *B. infantis* among all children in COPSAC$_{2010}$ cohort, children with and without breastfeeding at the stool sample collection age. Colour indicates the quartiles of SES index.

### Extended Data Table 1. Characterizations of CHILD samples.

| Category | Variable | Cohort |
|---|---|---|
| No. patients | | 3263 |
| Confounder | Male, n(%) | |
| | | 1717 (52.6%) |
| | Study center, n(%) | |
| | Edmonton | 768 (23.5%) |
| | Toronto | 768 (23.5%) |
| | Vancouver | 735 (22.5%) |
| | Winnipeg | 992 (30.4%) |
| Health | Atopy at 5y, n(%) | |
| | | 494 (19.3%) |
| | Unknown | 708 (21.7%) |
| | Asthma at 5y, n(%) | |
| | | 165 (6.9%) |
| | Unknown | 864 (26.5%) |
| | Overweight/obese at 5y, n(%) | |
| | | 542 (20.6%) |
| | Unknown | 638 (19.6%) |
| | Emotional/behavioral problems at 5y, n(%) | |
| | | 172 (7.3%) |
| | Unknown | 903 (27.7%) |
| Pregnancy | Maternal antibiotics (preg.), n(%) | |
| | | 314 (9.6%) |
| | Gestational diabetes, n(%) | |
| | | 154 (4.7%) |
| | Maternal distress (preg.) | |
| | Median (Range) | 13 (0, 35) |
| | IQR (Q1,Q3) | 9, 17 |
| | Unknown | 325 (10%) |
| | Prenatal smoking, n(%) | |
| | | 606 (19.1%) |
| | Unknown | 97 (3%) |
| Birth | Intrapartum antibiotics, n(%) | |
| | | 1326 (41.2%) |
| | Unknown | 45 (1.4%) |
| | Delivery mode, n(%) | |
| | Vaginal | 2412 (74.8%) |
| | C-section with labor | 425 (13.2%) |
| | C-section without labor | 387 (12%) |
| | Unknown | 39 (1.2%) |
| | Birth weight Z score | |
| | Median (Range) | -0.1 (-3.1, 4.3) |
| | IQR (Q1,Q3) | -0.7, 0.5 |
| | Unknown | 75 (2.3%) |
| | Gestational age | |
| | Median (Range) | 278 (238, 300) |
| | IQR (Q1,Q3) | 272, 283 |
| | Unknown | 54 (1.7%) |
| | Season of birth, n(%) | |
| | Spring | 889 (27.2%) |
| | Summer | 830 (25.4%) |
| | Fall | 754 (23.1%) |
| | Winter | 790 (24.2%) |
| Postnatal | Antibiotics use (1y), n(%) | |
| | | 619 (26.1%) |
| | Unknown | 890 (27.3%) |
| | Reg. childcare attend. (1y), n(%) | |
| | | 735 (28.5%) |
| | Unknown | 688 (21.1%) |
| | Reg. childcare attend. (3m), n(%) | |
| | | 338 (11.3%) |
| | Unknown | 274 (8.4%) |
| | Any breastfeeding dur. | |
| | Median (Range) | 10 (0, 30) |
| | IQR (Q1,Q3) | 5, 14 |
| | Unknown | 106 (3.2%) |
| | Pet (cat) (1y), n(%) | |
| | | 550 (21.4%) |
| | Unknown | 690 (21.1%) |
| | Pet (dog) (1y), n(%) | |
| | | 701 (27.1%) |
| | Unknown | 681 (20.9%) |
| | Older siblings (#) | |
| | Median (Range) | 0 (0, 4) |
| | IQR (Q1,Q3) | 0, 1 |
| | Unknown | 101 (3.1%) |
| | Postnatal smoking, n(%) | |
| | | 620 (23.1%) |

| | | |
|---|---|---|
| | Unknown | 578 (17.7%) |
| | Exclusive breastfeeding (6m), n(%) | |
| | | 522 (17.1%) |
| | Unknown | 208 (6.4%) |
| Home Environment | Cleaning product use | |
| | Median (Range) | 31 (4, 76) |
| | IQR (Q1,Q3) | 25, 38 |
| | Unknown | 341 (10.5%) |
| | Multifamily home, n(%) | |
| | | 878 (28.9%) |
| | Unknown | 223 (6.8%) |
| | Clutter, n(%) | |
| | | 581 (19.1%) |
| | Unknown | 229 (7%) |
| | Potted plant, n(%) | |
| | | 1822 (59.9%) |
| | Unknown | 223 (6.8%) |
| | Carpet floor, n(%) | |
| | | 1722 (56.6%) |
| | Unknown | 223 (6.8%) |
| | Medium/high mould, n(%) | |
| | | 359 (11.8%) |
| | Unknown | 223 (6.8%) |
| Neighbourhood Environment | Dwelling density | |
| | Median (Range) | 0.2 (-0.8, 10.7) |
| | IQR (Q1,Q3) | -0.2, 1.4 |
| | Unknown | 197 (6%) |
| | Air pollution (NO2) (preg.) | |
| | Median (Range) | 10.6 (1.3, 35.8) |
| | IQR (Q1,Q3) | 5.3, 15 |
| | Unknown | 191 (5.9%) |
| | Air pollution (NO2) (1y) | |
| | Median (Range) | 9.1 (0.5, 30.5) |
| | IQR (Q1,Q3) | 4.6, 13.3 |
| | Unknown | 194 (5.9%) |
| | Tree density (preg.) | |
| | Median (Range) | 20 (0, 99) |
| | IQR (Q1,Q3) | 13, 32 |
| | Unknown | 663 (20.3%) |
| | Rural, n(%) | |
| | | 186 (6%) |
| | Unknown | 146 (4.5%) |
| Parental Diet/Health | Paternal asthma, n(%) | |
| | | 510 (19.2%) |
| | Unknown | 600 (18.4%) |
| | Paternal atopy, n(%) | |
| | | 1663 (67.7%) |
| | Unknown | 806 (24.7%) |
| | Maternal asthma, n(%) | |
| | | 749 (23.4%) |
| | Unknown | 56 (1.7%) |
| | Maternal distress (1y) | |
| | Median (Range) | 12 (0, 40) |
| | IQR (Q1,Q3) | 7, 17 |
| | Unknown | 685 (21%) |
| | Maternal BMI | |
| | Median (Range) | 23.3 (13.9, 56.9) |
| | IQR (Q1,Q3) | 21, 27 |
| | Unknown | 327 (10%) |
| | Maternal UPF diet | |
| | Median (Range) | 46.8 (1.9, 82.9) |
| | IQR (Q1,Q3) | 39.7, 53.8 |
| | Unknown | 291 (8.9%) |
| | Maternal atopy, n(%) | |
| | | 1727 (57.7%) |
| | Unknown | 268 (8.2%) |

**Extended Data Table 2. Prevalence of Bifidobacterium species in children breastfed up to 3 months across CHILD study sites and $COPSAC_{2010}$ cohort.** q-value is calculated based on chi-square test and with adjustment of multiple comparisons using Benjamini-Hochberg (BH) method.

| Species | Vancouver | Toronto | Winnipeg | Edmonton | q-value across CHILD study sites | CHILD Study | $COPSAC_{2010}$ Cohort | q-value between CHILD study and $COPSAC_{2010}$ cohort |
|---|---|---|---|---|---|---|---|---|
| *Bifidobacterium adolescentis* | 28% | 24% | 33% | 29% | 0.12 | 29% | 83% | <0.001 |
| *Bifidobacterium animalis* | 44% | 51% | 47% | 48% | 0.47 | 47% | <10% | <0.001 |
| *Bifidobacterium bifidum* | 48% | 40% | 50% | 45% | 0.12 | 46% | 89% | <0.001 |
| *Bifidobacterium breve* | 66% | 60% | 56% | 63% | 0.12 | 61% | 94% | <0.001 |
| *Bifidobacterium dentium* | 36% | 40% | 39% | 34% | 0.47 | 38% | 38% | 0.94 |
| *Bifidobacterium infantis* | 37% | 30% | 23% | 17% | <0.001 | 28% | 75% | <0.001 |
| *Bifidobacterium longum* | 89% | 87% | 87% | 89% | 0.94 | 88% | 100% | <0.001 |
| *Bifidobacterium pseudocatenulatum* | 31% | 34% | 36% | 38% | 0.47 | 35% | 86% | <0.001 |
